# Economic Impact Payment, Human Mobility and the COVID-19 Mitigation in the United States

**DOI:** 10.1101/2021.05.17.21257358

**Authors:** Ruohao Zhang

## Abstract

This paper studies the relationship between the individual’s income and COVID-19 mitigation effort contribution. The paper suggests that in addition to the government mandatory policies, the income compensation policy is an alternative government instrument that helps increase the individual and social aggregate COVID-19 mitigation effort. I empirically test the effect of the income compensation policy by utilizing the United States economic impact payment (EIP) in April 2020 as a quasi-natural experiment, and use the cellphone GPS measured human mobility data as the outcome indicator of the COVID-19 mitigation effort. I find that by receiving EIP, individuals on average significantly increase median home dwell time by 3% – 5% (about 26–45 minutes). This paper highlights an unintended effect of EIP: in addition to providing economic assistance, EIP also helps mitigate the COVID-19 virus transmission.

**JEL codes:** D11 D12 H41 I12 I14 I18

## 1 Introduction

In the absence of a medical countermeasure, the non-pharmaceutical interventions (NPI) have become the main tool to mitigate the COVID-19 pandemic in 2020. Under the NPI, the public is expected to devote the mitigation effort, such as social distancing, wearing masks, shelterin-place and quarantine. Although the mitigation effort made by the uninfected people only protects themselves from being infected, those made by the infected people can protect the others. Due to the limited testing resources and similar symptoms as influenza, in the early stage of COVID-19 pandemic, the majority population do not know whether they are infected or not. This information gap creates an incentive for all individuals, regardless of their infection status, to contribute the mitigation effort. The mitigation effort slows down the spreading of the virus in the society, so that it can be treated as a public good.

Due to the nature of the public good, the market determined individuals’ contribution to the mitigation effort is always lower than the social optimal level. The government interventions help increase the social aggregate mitigation effort and raise the associated social welfare. A typical instrument of the government interventions is the mandatory policies, which can raise the lower bound of the mitigation effort contribution for all individuals. If properly enforced, the mandatory policies could be very effective in raising the social aggregate mitigation effort. However, in many cases the mandatory policies are difficult to be enforced due to varies of social and politic issues. In this paper, I argue that other than the mandatory policies, the income compensation is a second policy instrument that is available for the government to increase the social aggregate mitigation effort. The income compensation, as a policy instrument, gets much less attention but could be a good complement to the mandatory policies. According to the public goods theory, if the income compensation is properly customized for each individual based on her income and the status quo social aggregate mitigation effort, then it can be effective in raising the individual’s optimal level of mitigation effort so that increasing her mitigation effort contribution.

I use a data set of human mobility to empirically test the argument that the income compensation may raise the mitigation effort contribution, because the decrease in the human mobility is a typical mitigation effort during the early stage of the pandemic. I utilize the first round economic impact payment (EIP) in April, 2020 as a quasi-natural experiment. EIP, which is an exogenous payment to the individuals in the United States for economic assistance during the COVID-19 pandemic, provides an opportunity to test whether there is a change in individual mobility in response to the income compensation. The empirical results are highly consistent with the argument, suggesting that EIP caused a 3% to 5% (about 26–45 minutes) increase in daily home dwell time. Among all the income groups, only the mobility of the households with income less than $20,000 appear to be unaffected by EIP. This is because for these households, the status quo social aggregate mitigation effort is higher than their optimal mitigation effort, and EIP amount is not large enough to raise their optimal mitigation effort above the social aggregate level, so they have no incentive to contribute additional mitigation effort both before and after receiving EIP. For all the other income groups, receiving EIP increases their daily home dwell time, suggesting that EIP motivates them to increase contribution to the mitigation effort. In addition, the results also suggest that the government mitigation policies are not strictly enforced in the United States, so that there is no mandatory mitigation requirement.

This paper contributes to three bodies of literature. Firstly, the previous public goods literature has illustrated the relationship between income and the contribution to public goods (Buckley and Croson, 2006; Kotchen and Moore, 2007; Jacobsen et al., 2012). I expand the literature by enhancing the public goods model in Kotchen and Moore (2007) to include a potential mandatory requirement of public good contribution. Besides, I also derive the heterogeneous changes in the contribution to public good in response to the income compensation for the individuals with different income levels. Secondly, the empirical literature of the economic impact payments and the other similar stimulus payments during the past crises in the United States, such as 2001 tax rebate and 2008 stimulus payment, has shown that the stimulus payments significantly increased the household private consumption (Johnson et al., 2006), but have adverse health impacts, possibly because of the increase in alcohol and drug consumption (Evans and Moore, 2011; Gross and Tobacman, 2014). I contribute to this literature by focusing on a different consumption category, the public good consumption. Last but not least, in the current and rapidly evolving literature related to COVID-19, there are several papers studying the effects of the government policies during the pandemic on human mobility (Dave et al., 2020; Gupta et al., 2020; Wright et al., 2020), and the relationship between human behavior and the spread of the coronavirus (Farboodi et al., 2020; Chudik et al., 2020; Fowler et al., 2020; Kapoor et al., 2020). There are other empirical studies focusing on the different level of the compliance to the government mitigation policies (Gollwitzer et al., 2020; Allcott et al., 2020; Wright et al., 2020). However, to the best of my knowledge, this is the only paper studying the public goods nature of the COVID-19 mitigation effort, and inferring a possible decentralized solution that increases the individual and social mitigation effort. The paper points out that in addition to its original purpose, EIP, or the stimulus check, also motivates individuals to increase their contribution to the COVID-19 mitigation effort.

## 2 Background and Testable Hypothesis

### 2.1 Theoretical Background

In the context of a global infectious disease outbreak, the mitigation effort can be treated as a consumable good because it brings private utility gains through reducing the risk of being infected. Consuming mitigation effort also yields private cost including the cost of less working hours, personal protection equipment purchase, restricted travel plans, cancellation of entertainment events and so on. At the same time, the mitigation effort can be treated as a public good because it reduces the virus transmission rate and decreases the infection risk for the public.

As a public good, the social aggregate mitigation effort is non-rival and non-excludable. Each individual determines her own optimal mitigation effort based on her income and all the other individuals’ optimal mitigation effort in the society. Social equilibrium can be achieved when the social aggregate mitigation effort is greater or equal to everyone’s optimal mitigation effort. Given the assumption that the mitigation effort is a normal good, there exists an income threshold level set by the status quo social aggregate mitigation effort at the equilibrium, which determines whether an individual is a “voluntary contributor” of the mitigation effort or not.^1^ Only the individuals with income above the threshold are defined as “voluntary contributors”, because their optimal mitigation effort is higher than the aggregate mitigation effort contributed by all the others in the society, so that they contribute additional positive mitigation effort to raise the social aggregate level. For the others with income below the threshold, they are defined as “non-voluntary contributors”, because their optimal mitigation effort is lower than the aggregate mitigation effort contributed by the others, so that they have no incentive to raise social aggregate level and contribute zero mitigation effort (unless there are mandatory requirements). In Appendix A.1, I describe the detail of a formal theoretical model that illustrates the individual’s behavior in contributing the mitigation effort at the social equilibrium.

The market determined mitigation effort contribution is always insufficient for social optimal solution due to its public good nature and the positive externalities, suggesting that the government interventions are desired to increase the social aggregate mitigation effort contribution. One commonly used intervention instrument is the mandatory policies. Under the government mandatory policies, the individual mitigation effort consists of two parts: the mandatory required mitigation effort and the additional voluntary mitigation effort. The mandatory required mitigation effort is non-negative and serves as a lower bound of the individual mitigation effort. The individuals who are “non-voluntary contributors” only contribute at the mandatory level, whereas “voluntary contributors” contribute positive additional voluntary mitigation effort other than the mandatory requirement.

In addition to the mandatory polices, there is a second government intervention instrument that we are less aware of: the compensation on individual income. An income compensation may be effective in increasing the social aggregate mitigation effort contribution in two ways: For the individuals who are “voluntary contributors”, recall that their income is above the threshold and their ex-ante additional voluntary mitigation effort is positive, so that even a small amount of compensation that marginally increase their income will increase their additional voluntary mitigation effort. However, for the individuals who are “non-voluntary contributors”, they contribute at mandatory level (contribute zero if no mandatory requirement) as long as their income is below the threshold. A marginal compensation do not affect their mitigation effort contribution. Instead, the compensation must be large enough to raise their income above the threshold therefore covert them to the “voluntary contributors” and make positive additional voluntary mitigation effort.

This theory has important policy implications. The government can increase the social aggregate mitigation effort through the mandatory policies. However, due to politic and social issues, sometimes it is difficult for the government to enforce the mandatory policies or increase the stringency of the existing mandatory policies. As an alternative, the income compensation, if properly distributed, can be a good complement to the mandatory policies. In order to be effective, the income compensation should be carefully customized for “non-voluntary contributors” with ex-ante income lower than the threshold, so that it can help raise their income above the threshold and increase their mitigation effort contribution.

### 2.2 Empirical Background

On March 27th, 2020, the Coronavirus Aid, Relief, and Economic Security (CARES) Act was passed by the U.S. Congress and signed into law by President Trump. The CARES Act contains a $2 trillion economic relief package that aims to provide assistance for the households, small businesses, the healthcare system and local governments.^2^ A portion of the $2 trillion package is the economic impact payment (EIP) to the households as economic assistance during the pandemic. The eligible taxpayer, who is single and with annual income less than $75,000, or the head of a household with annual income less than $112,500, receives $1,200 EIP. The eligible married couple that files taxes jointly with annual income less than $150,000 receives $2,400 EIP. A household with children under 17 years old receives additional $500 for each child up to 2 children. Taxpayers with higher annual income may also receive a reduced EIP. The first wave of EIP distribution was made by the IRS starting in the week of April 13th, 2020 through direct deposits, followed by the subsequent paper check distribution.^3^ Although the primary purpose of EIP is to provide economic relief, this lump-sum payment also exogenously increase the recipients’ income, so that it can be used to test the income effect the COVID-19 mitigation effort contribution.

Recall that the individual contribution of the COVID-19 mitigation effort consists of two components: the mandatory mitigation effort enforced by the government policies, and the additional voluntary mitigation effort made by the individuals. The mandatory contribution level is the minimum contribution that is strictly enforced by the government. If there is a policy that is not effectively enforced and relies on self-compliance, then I consider the associated mitigation effort as the additional voluntary mitigation effort, since the level of compliance is voluntarily determined. In the United States, most COVID-19 NPIs are not being strictly enforced and heavily rely on self-compliance. That makes it hard to tell whether there is a meaningful mandatory contribution requirement. There are several papers studying the NPI effects on human mobility, providing strong evidences suggesting that the mitigation effort in the United States is mostly voluntary based. For example, Wright et al. (2020) find that under the shelter-in-place policy, the high income counties significantly reduce social mobility whereas nothing changes in the low income counties. The compliance to the NPIs also highly depends on various individual characteristics including partisanship and conservative media exposure (Allcott et al., 2020; Gollwitzer et al., 2020), social capital (Barrios et al., 2021), trust to the media, science and government (Bai et al., 2020; Barrios et al., 2021; Durante et al., 2020) and so on. Similar empirical results are found in this paper, showing mitigation policies is not effectively enforced, suggesting the nature of voluntary contribution of the mitigation effort in the United States.

### 2.3 Hypothesis

As an income compensation, EIP may affect the social aggregate mitigation effort contribution. In order to increase the mitigation effort contribution, the individuals who receive EIP must have their income that is *greater* than the corresponding income threshold (determined by the status quo social aggregate level) subtracted by the amount of EIP. These individuals are either “voluntary contributors” (if income is *greater* than the threshold), who will increase their migration effort in response to a marginal increase in income; or “non-voluntary contributors” (if income is *smaller* than the threshold but *greater* than the threshold minus EIP), so that they are converted to “voluntary contributors” after receiving EIP, and make positive additional voluntary mitigation effort. In both cases EIP increases these individuals’ mitigation effort contribution, which leads to higher social aggregate mitigation effort.

In the empirical analysis, I use human mobility at the census block group (CBG) level as an indicator of the COVID-19 mitigation effort. By assuming that in most CBGs, there are individuals with sufficient income level so that they will increase their mitigation effort contribution in response to EIP, we have the following hypothesis:

#### Main Hypothesis

*The aggregate human mobility in the CBG decreases in response to receiving EIP.*

There is another implication that is potentially testable: when the government raises the mandatory contribution level, then we should expect an increase in the aggregate mitigation effort contribution, mainly driven by the increase in the contribution from the lower income individuals to meet the mandatory requirement. In this paper, I do not focus on testing this hypothesis, because as discussed above, the COVID-19 NPIs in the United States are not strict enough to offer an empirical test. In the empirical analysis shown below, there is a lack of evidence supporting the argument that the government NPIs bind human mobility in the United States.^4^

## 3 Data

To empirically test the main hypothesis that EIP causes the decrease in human mobility, I assemble a CBG-by-day data set that covers 216,069 CBGs across 50 states and Washington D.C. in the United States (including Alaska and Hawaii). The sample period is between April 1st, 2020 and April 20th, 2020. The data set includes information on the household income distribution and daily human mobility at the CBG level, daily COVID-19 confirmed case and death numbers at the county level, and the government policies regarding the mandatory NPIs at both the state and county level.^5^

I obtain human mobility data from the “Social Distancing Metrics” database provided by SafeGraph, Inc. The data is collected from the GPS locations of 45 million smartphone devices in the United States and its territories, and aggregate at the CBG level. I use “*Median Home Dwell Time*” as the outcome variable to indicate human mobility. “*Median Home Dwell Time*” calculates the median minutes that the smart phone devices are at home through a single day. “Home” is identified as an 153*m* ×153*m* square that the smart phone devices are mostly located during the overnight hours between 6:00 PM and 7:00 AM. “Home” location is updated at the start of each month, according to the previous 6 weeks overnight hours locations. Safegraph forms a “confidence score” for the cell phones’ overnight hours locations. Devices with low confidence score of home location were treated as if the home location is unknown.^6^ One concern of this data is that the “Home” location may be misspecified for college students returning back home when the schools are closed. Since the analysis is limited within April, all cell phones in the sample have their “Home” location updated at April 1st, whereas school closures happened in mid March (Mangrum and Niekamp, 2020). Therefore, the college students leaving campus are treated as home location unknown, and do not affect the analysis.

The county level COVID-19 confirmed case load and death data (JHU data) are collected by Center for Systems Science and Engineering at John Hopkins University. JHU data is publicly available online, covering the whole time period of COVID-19 pandemic in the United States from January 22nd, 2020 (first case in U.S.) to today.^7^ The state and county level government NPI policy data (Keystone data) is obtained from Keystone Strategy.^8^ Keystone data is updated on April 16th, 2020, which collects information from government websites and local news reports. Keystone data contains the state level policies for all US states and territories, and the county level policies for all counties with at least 100 confirmed cases as of April 6th, 2020. Among all state and county policies reported by the Keystone data, I consider the following three policies that tend to have the most significant effect on human mobility: “Social Distancing”, “Non-essential Services Closure”, and “Shelter-in-Place”. “Social Distancing” is a requirement that people maintain physical distance and reduce the number of times people contact each other. “Non-essential Services Closure” pauses all the unnecessary economic activities. “Shelter-in-Place” is a more general and stricter order that requires people to minimize their social activities.^9^

Census data provide detailed information about the CBG characteristics. Since the census data do not have time variation on a daily basis, most of the characteristics are captured by the census block group fixed effect. However, the income distribution is important for two reasons. First, it identifies the percentage of the households receiving EIP according to the eligibility standard. Second, EIP effect on human mobility tends to be heterogeneous across income levels. The EIP eligibility is based on 2018 or 2019 annual income reflected on the tax file. Census only reports income at household unit, and the 2019 income data at the CBG level is not available yet, so I use Census 2014-2018 five year estimates household income data to proxy the current income distribution.^10^

Table 1 reports the summary statistics. The first panel of the table reports median home dwell time, percentage of the households in each income group and the county COVID-19 statistics. On average, the median home dwell time is 886 minutes (around 15 hours) per day. About 87.59% of all the households in each CBG have income lower than $150,000. The second panel reports the state and county policies. It shows that only 13 states issued “Non-essential Services Closure” order, because most of the states skipped it and directly issued the more strict “Shelter-in-Place” order. These 13 states either issue the “Shelter-in-Place” order after “Non-essential Services Closure” order, or they have not formally issued a “Shelter-in-Place” order yet. The county level policies can be different from the state level policies since many county governments move faster than the state and set stricter policies.^11^

**Table 1:**
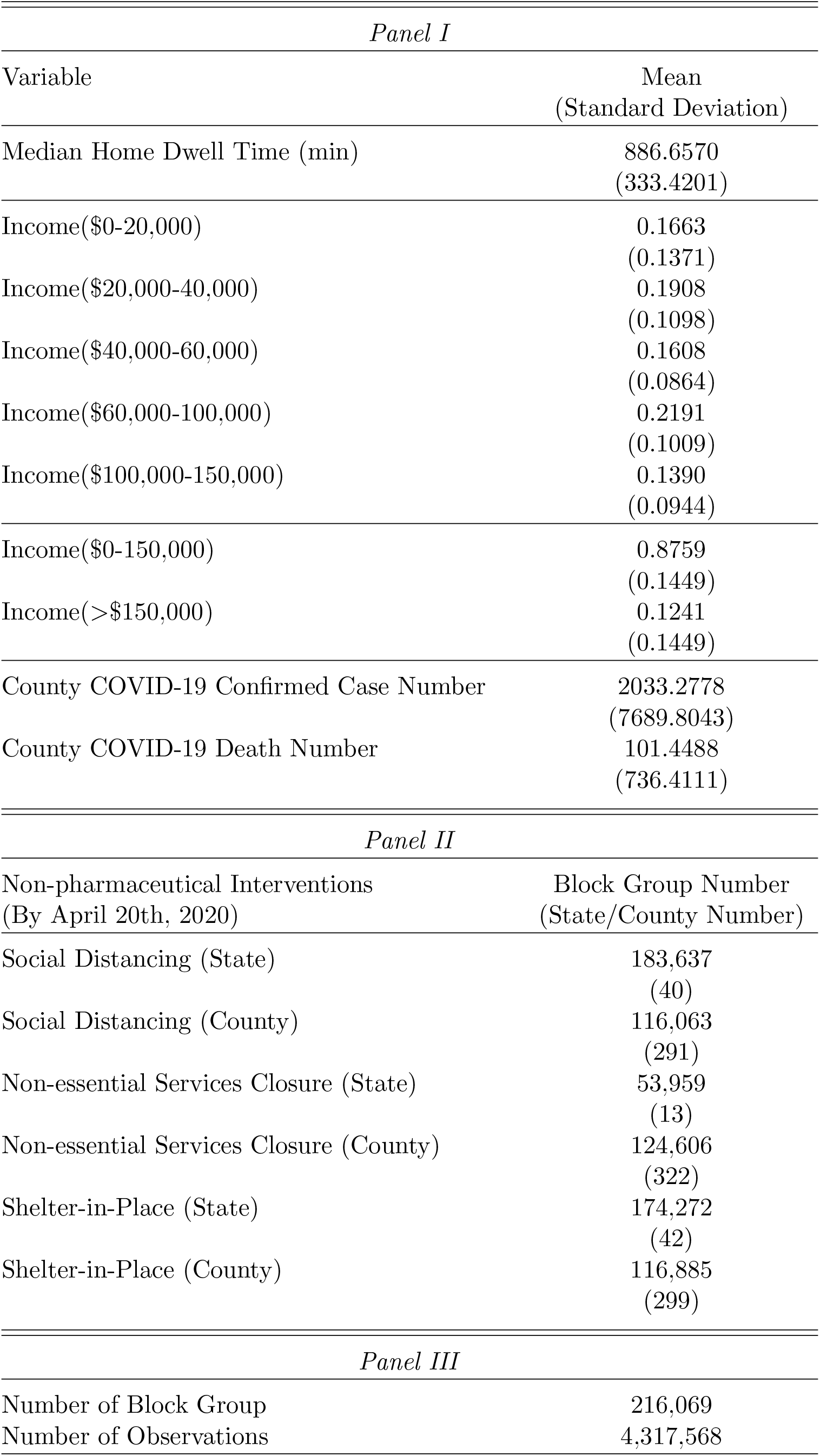
Descriptive Statistics

Figure 1 shows the unconditional average daily trend of “*Median Home Dwell Time*” across all CBGs in the sample. As expected, in general “*Median Home Dwell Time*” is lower during the weekdays and higher during the weekends. By comparing across weeks, the figure shows that for the week when EIP is distributed, there is much higher “*Median Home Dwell Time*”, suggesting people on average increase their home dwell time in that week.

**Figure 1:**
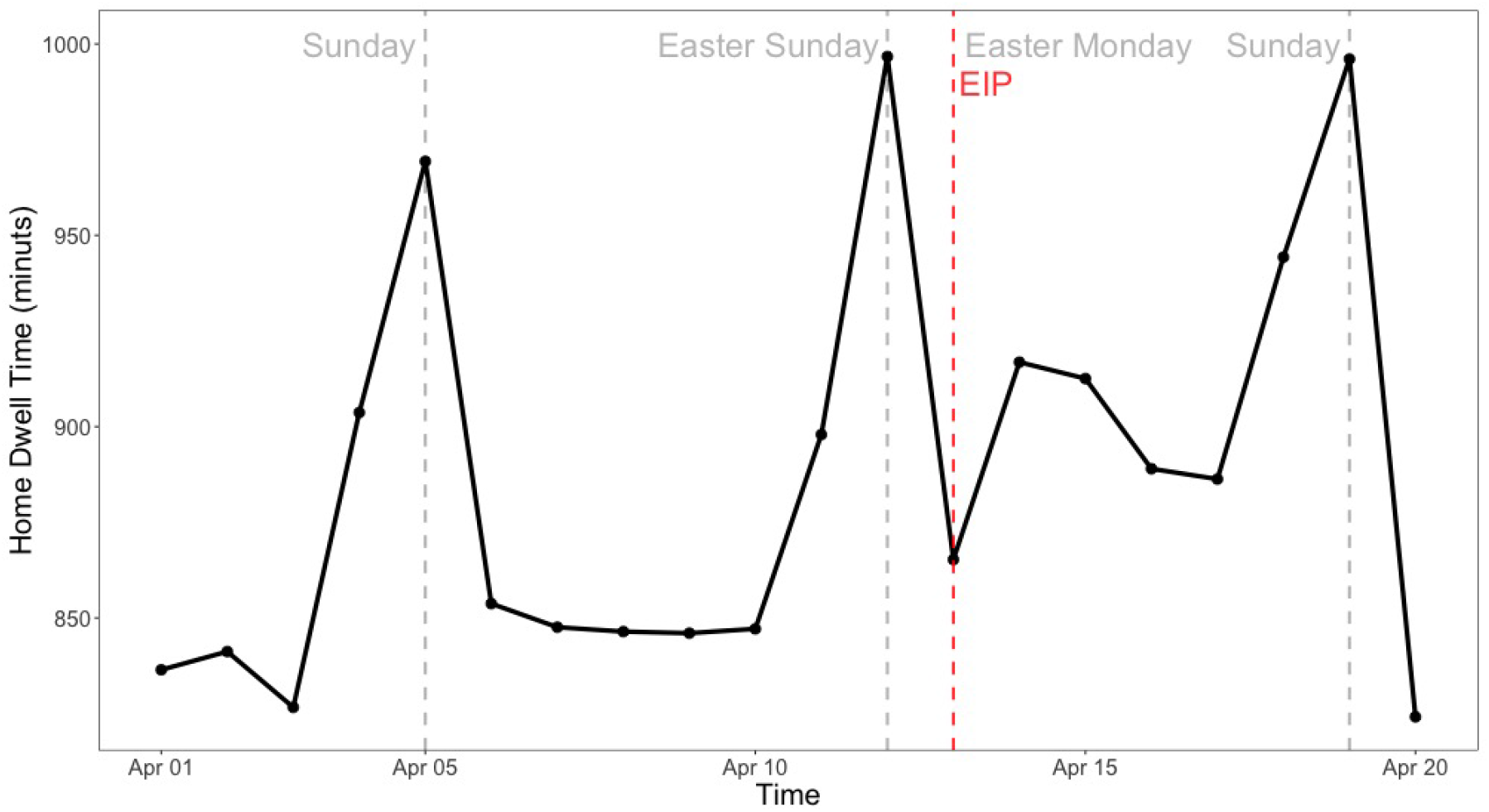
Daily average Median Home Dwell Time

## 4 Empirical Analysis

### 4.1 An Overview of Identification Strategies

In the empirical analysis, the outcome variable is the CBG level median mobility that measures the COVID-19 mitigation effort. The treatment is EIP, which is exogenous to the individuals. According to the hypothesis that EIP decreases the human mobility, I expect EIP to increase mitigation effort contribution (lower mobility) as it compensates the individuals’ income. One major challenge of the analysis is that EIP is a (near) universal treatment for all CBGs across the whole United States, so that we do not have a control group of the CBGs that are not treated by EIP. This is because within almost all CBGs there are households that are eligible for and have received EIP. However, the EIP treatment on each CBG is different, depending on the ratio of the households who are eligible for EIP. I utilize the variation in the EIP treatment sizes across CBGs to identify the EIP treatment effect on human mobility in two different study designs: an event study that compares two groups of CBGs that receive different sizes of the EIP treatment, and a difference-in-differences framework (DID) with continuous variation in the EIP treatment.

The event study compares the changes in human mobility before and after the EIP treatment on a daily basis, which illustrates the causal effect of the EIP treatment intuitively. However, it does not use all the treatment variation. Instead, the event study suppresses the continuous variation in the EIP treatment in a binary setting by comparing two arbitrarily defined groups of CBGs. The DID with continuous treatment is an extension of the classic binary potential outcome framework, which identifies the average treatment effect by allowing a series of potential outcomes for the CBGs with continuous variation in the treatment level. However, it is challenging to test the common trend assumptions for the DID with continuous treatment since there is no clear distinction between treatment group and control group. Since both methods have their own pros and cons, they complement each other to provide more convincing empirical analysis.

I conduct all the analysis using both state and county level policy data. Compared with the analysis using state level policy data, the county level policy data captures the potential mandatory mitigation requirement more precisely. In addition, it also allow me to test if the EIP effect is different in the outbreak counties with more than 100 confirmed cases. An implicit assumption in the empirical analysis is that the household ex-ante income (excluding EIP) is fixed across the study period. This is a relatively strong assumption, since there is a large wave of lay-offs/furloughs during the pandemic. Although the day fixed effect captures the average income trend across all CBGs, with the possibility that lower income workers are more likely to be laid off/furloughed, the variation in contemporary income shocks from unemployment (because of the different income distributions across the CBGs) may still causes bias and failure to establish a causal relationship. I address this problem in two ways. First, I use a short study period between April 1st, 2020 and April 20th, 2020. According to BLS (2020) and Coibion et al. (2020), the major wave of lay-offs occurred before the beginning of April, so the income shocks due to lay-offs/furloughs is mild through out the study period.^12^ Second, in the DID framework, I conduct robustness check analysis with a weaker time trend assumption, which allow state and county specific time fixed effects. These relaxed assumptions can partially capture the unobserved variation in income shocks from unemployment at the state/county level. Therefore, if there are heterogeneous time trends across the states/counties due to unobserved variables, then the state/county specific time fixed effects will improve the estimation consistency. I find the state/county specific time fixed effects do not change the coefficients, suggesting that the assumption of constant income level (detrended by the day fixed effects) does not bias the treatment effect nor violate the causal inference.

### 4.2 Event Study

I set an arbitrarily defined cutoff of the ratio of the households with income less than $150,000 (who are eligible for receiving EIP), and use the cutoff to separate all CBGs into two groups: the high income CBGs and the low income CBGs.^13^ The high income CBGs have the EIP eligible households ratio below the cutoff, and the low income CBGs have the ratio above the cutoff. By this definition, the high income CBGs receive relatively less EIP treatment than the low income CBGs. The event study can detect the EIP treatment effect by estimating the differences in human mobility between the high and low income CBGs on a daily basis and comparing these daily differences before and after the EIP treatment. According to the main hypothesis that EIP reduces the human mobility, I expect to observe significant changes in the daily differences after the EIP treatment, due to the different size of the EIP treatment received by each group of CBGs. I set the cutoff at 75%, and compare both the unconditional and conditional daily trend of “Median Home Dwell Time” between two groups of CBGs.^14^

Figure 2 shows the unconditional average daily gap of “*Median Home Dwell Time*” between the high and low income CBGs, where the reference group is the high income CBGs. As we expected, the gap is always negative, suggesting the low income CBGs always has lower “*Median Home Dwell Time*” than the high income CBGs. It is worth to mention that although there are some pre-existing weekday versus weekend variation in the gap, after EIP is received, the gap decreases in both weekdays and weekends. I follow the same definition in Figure 2 to compare the conditional daily gap in “Median Home Dwell Time” between low and high income CBGs.

**Figure 2:**
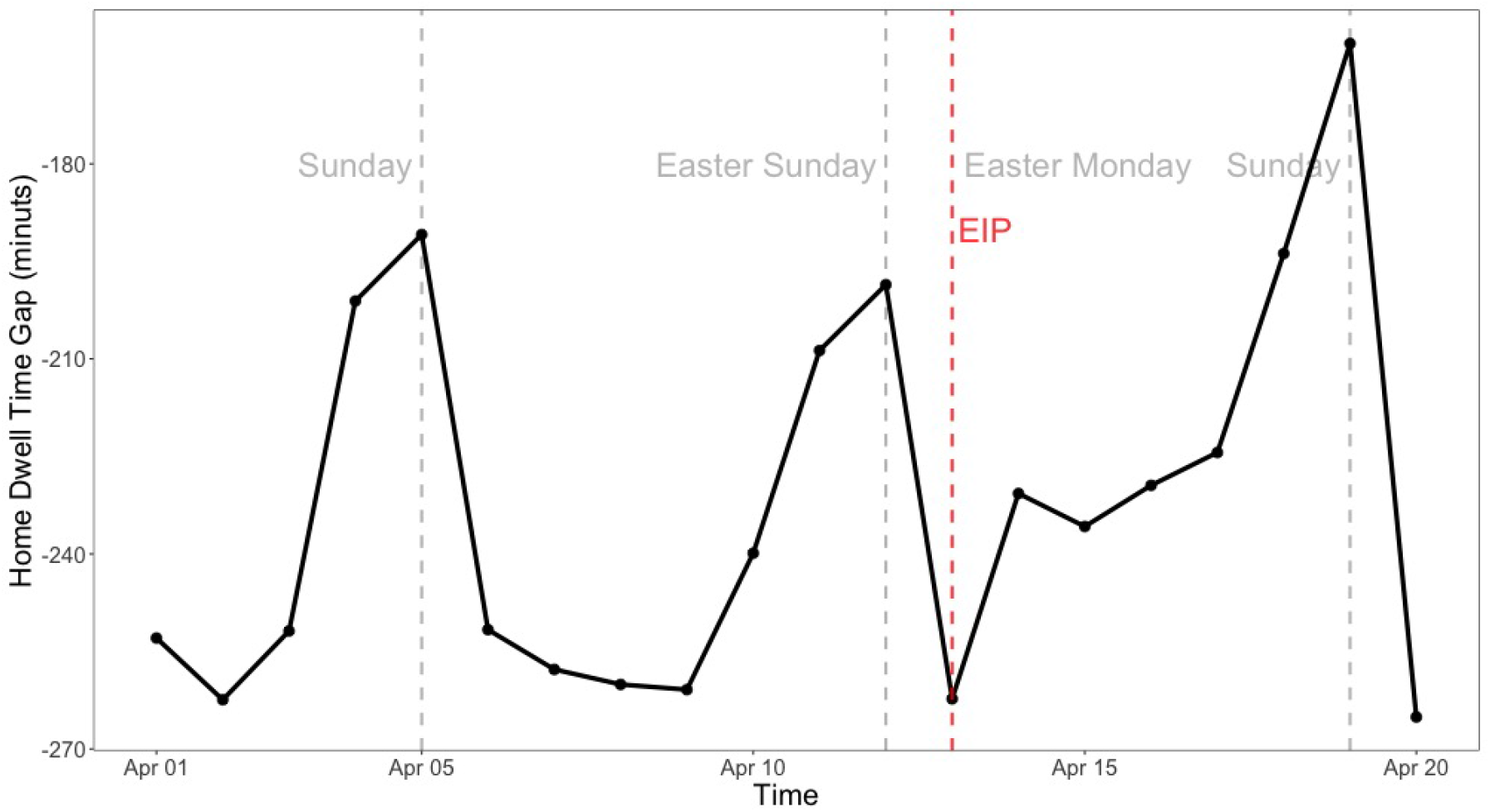
Daily Median Home Dwell Time gap between CBGs with share of EIP reception more/less than 75%

Consider the following event study regression:

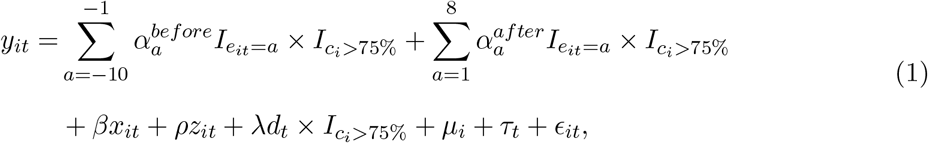

where *i* is the CBG index, *t* is the date index. *y*_*it*_ is the CBG human mobility, *x*_*it*_ is the covariates which capture the time-varying characteristics that are correlated with the human mobility. I define *x*_*it*_ to be the number of COVID-19 confirmed cases and deaths at county level because they are correlated with the time-varying individual preferences. *z*_*it*_ is a vector of three binary variables indicating different state/county level government policies, which captures the potential mandatory required mitigation effort contribution. *µ*_*i*_ and *τ*_*t*_ are the CBG and date fixed effects. *e*_*it*_ is the difference in days between date *t* and April 12th, 2020 (one day before receiving EIP). *c*_*i*_ is the share of the households eligible for receiving EIP (income less than $150,000).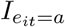 and 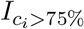 are indicator functions, which equal 1 if the conditions in the subscripts are satisfied and 0 otherwise. *d*_*t*_ is a dummy variable with *d*_*t*_ = 0 if date *t* is a weekday and *d*_*t*_ = 1 if date *t* is Saturday or Sunday. Since Figure 2 suggests that the gap vary across weekdays and weekends regardless of the EIP treatment, 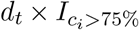 captures that preexisting variation. 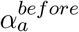 and 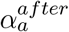 capture the daily gap between the high and low income CBGs.

I estimate the regression equation 1 using both the state and county level policy data, and plot the estimated daily gap 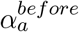 and 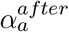. There is a dramatic difference in sample size between the state and county level policy data: the state level policy data contain 4,317,586 observations, the county level policy data contain 2,541,943 observations. This is because the county policy data only include counties with more than 100 cases as of April 6th, 2020. Figure 3 and 4 show the results with state and county policies respectively. Despite the big difference in the sample size, both figures show similar results: before the EIP treatment, the daily gaps are mostly negative; after the EIP treatment, there is a sharp increase in daily gaps, and such change persists for about one week.

**Figure 3:**
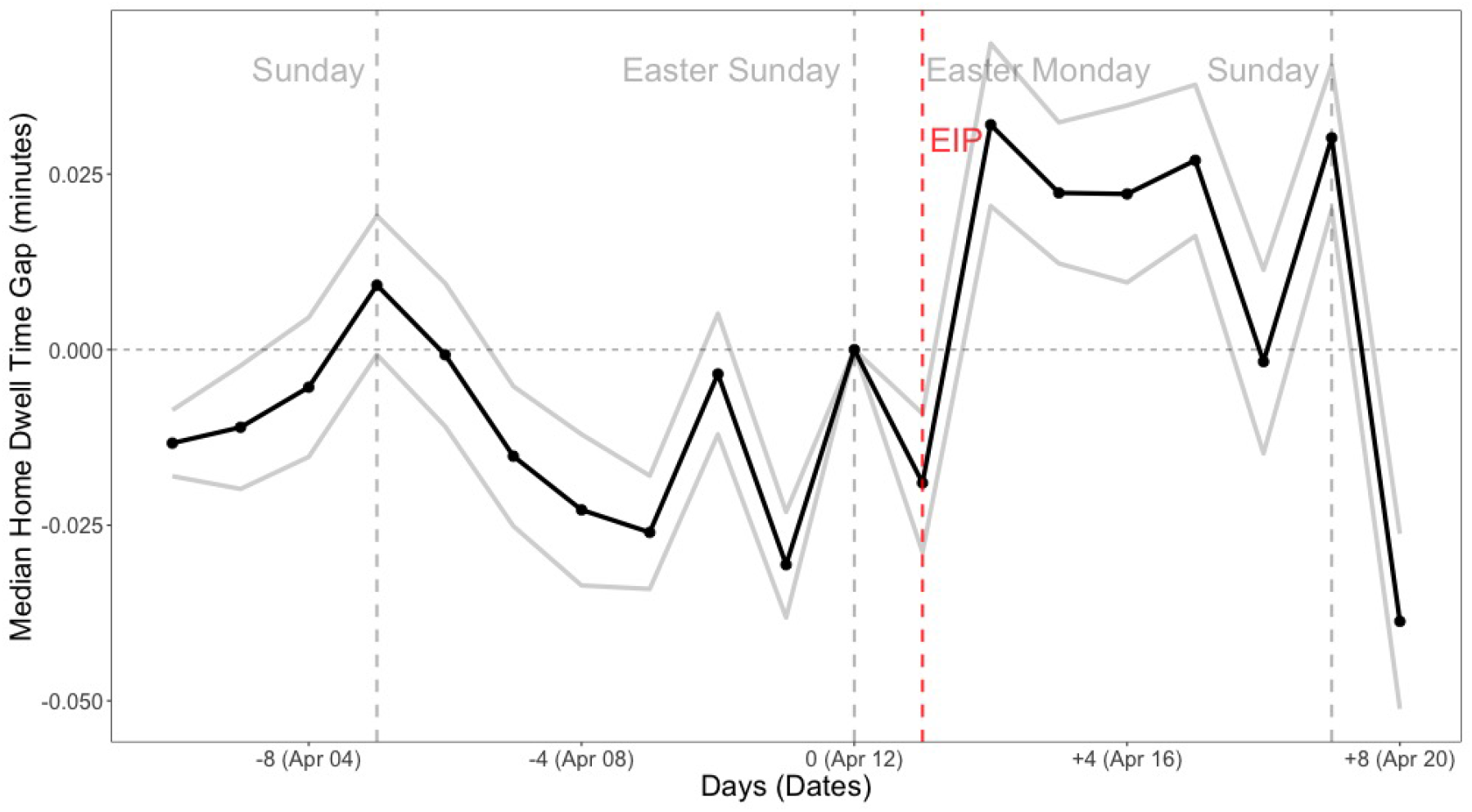
Event Study, State Policies

**Figure 4:**
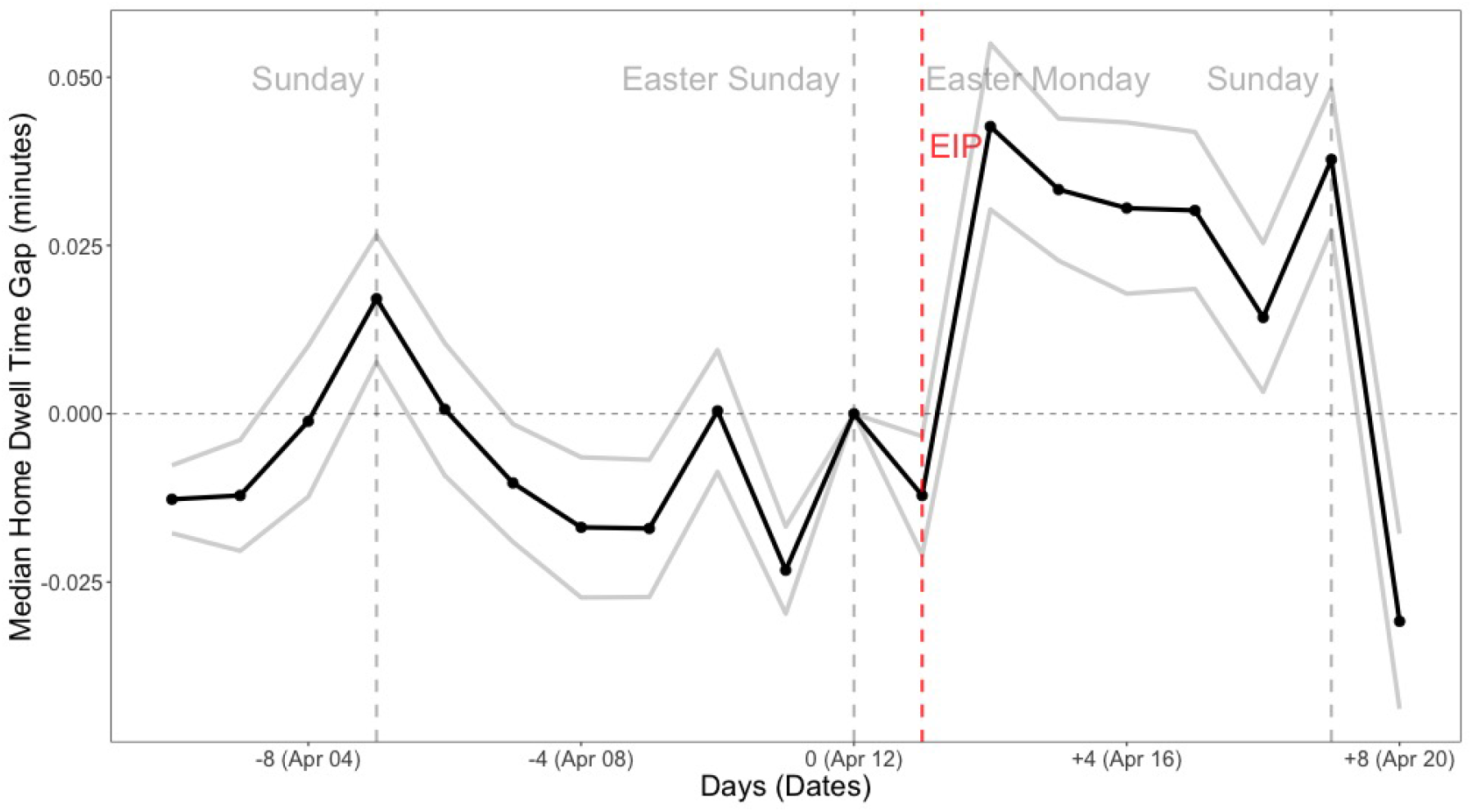
Event Study, County Policies

These results illustrate a causal relationship between the EIP treatment and the mitigation effort (human mobility), suggesting that compared with the high income CBGs, after receiving EIP, the “Median Home Dwell Time” in the low income CBGs increases dramatically, because the low income CBGs receive larger EIP treatment than the high income CBGs. These results are consistent with the main hypothesis that the EIP treatment decreases the human mobility.

### 4.3 Difference-in-Differences with Continuous Treatment

In the DID with continuous treatment setting, the continuous treatment is constructed based on the ratio of the households who are eligible for and have received EIP. Consider the following model:

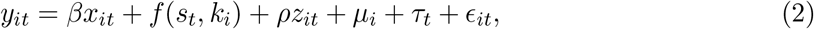

*f* (*s*_*t*_, *k*_*i*_) is a continuous function representing the EIP treatment received by the CBG *i* on date *t. s*_*t*_ is a dummy variable with *s*_*t*_ = 1 representing EIP has been received. *k*_*i*_ is the time invariant CBG income distribution that determines EIP eligibility.

*f* (·) is an unknown functional form of the continuous treatment, or a “dose-response” function of receiving different amounts of EIP.^15^ I define *s*_*t*_ = 1 if *t* is April 13th, 2020 or later. *k*_*i*_ is the percentage of the households with income less than $150,000 in the CBG *i*. I assume the functional form to be *f* (*s*_*t*_, *k*_*i*_) = *γ* × *s*_*t*_ × *k*_*i*_. Unlike a standard DID model with clear distinction between the treatment and control groups, almost all CBGs receive the EIP treatment, thus the treatment variation appears in the different amounts of EIP received across CBGs, which allows me to assess the treatment effect as a dose response.^16^ Therefore, instead of directly estimating the average treatment effect, this model specification infers the average treatment effect by making a functional form assumption for *f* (·) and estimating the average marginal effect (“average dose-response”) of receiving different amount of EIP. The sample average treatment effect can be identified in a DID setting by estimating the coefficient *γ*.^17^

Similar as the event study, I conduct the DID analysis using both state and county level policy data. Table 2 shows the main results. I find that the coefficients obtained from the two samples are identical in terms of signs and significance, and have very similar magnitudes. The results show that after receiving EIP, a CBG with a larger fraction of the households eligible for EIP has higher “*Median Home Dwell Time*”. The effect of the EIP treatment on “*Median Home Dwell Time*” is between 3% and 5.5%. This coefficient can be interpreted as the percentage increase in the CBG “*Median Home Dwell Time*” when all the households in the CBG are eligible for EIP.^18^ The coefficients on the other covariates are mostly insignificant, suggesting that the CBG and date fixed effects have captured most of the variations apart from the EIP treatment effect.^19^ The insignificant coefficients on the government policies may driven by the fact that the government policies are not effectively enforced.^20^ The magnitude of the coefficients on “County COVID-19 Confirmed Case” and “County COVID-19 Death” indicates small conditional correlation between the number of COVID-19 confirmed cases and deaths and human mobility. This result is in line with the previous work (Gupta et al., 2020; Engle et al., 2020) that finds such conditional correlation only exists at the early stage of the pandemic (in early March). In general, these results are consistent with the main hypothesis that the EIP treatment decreases human mobility.

**Table 2:** Main Result

|  | *Dependent variable:* |  |
| --- | --- | --- |
|  | *Median Home Dwell Time (minutes, in log)* |  |
|  | (State Policy) | (County Policy) |
| Economic Impact Payment | 0.0309***<br>(0.0108) | 0.0550***<br>(0.0091) |
| County COVID-19 Confirmed Case (per 100) | 0.0003*<br>(0.0002) | 0.0002<br>(0.0002) |
| County COVID-19 Death (per 100) | −0.0041***<br>(0.0012) | −0.0034***<br>(0.0012) |
| Social Distancing (State/County) | 0.0097*<br>(0.0050) | 0.0138<br>(0.0096) |
| Non-essential Services Closure (State/County) | 0.0048<br>(0.0090) | −0.0129<br>(0.0146) |
| Shelter-in-Place (State/County) | 0.0033<br>(0.0089) | 0.0023<br>(0.0131) |
| Block Group FE | Y | Y |
| Date FE | Y | Y |
| Observations | 4,317,586 | 2,541,943 |
| R<sup>2</sup> | 0.8681 | 0.8769 |
| Adjusted R<sup>2</sup> | 0.8612 | 0.8704 |
| Residual Std. Error | 0.4010 (df = 4101492) | 0.4006 (df = 2414702) |

*Note:* \*p<0.1; \*\*p<0.05; \*\*\*p<0.01
*Standard errors are clustered at state level.*

There might be some alternative mechanisms that generate the positive EIP effect on “*Median Home Dwell Time*”. For example, it may caused by an increase in leisure due to higher income. However, it is unlikely that the leisure can be affected in 20 days period with a one-time $1,200 payment. Even if there is an effect, it tends to be shown on the relatively lower income households, because EIP raises their income in a relatively larger portion. In the following section, I discuss the heterogeneous EIP effect across different income groups, and the results show that EIP has zero effect on low income households, which suggests the results are not driven by the changes in leisure.

## 5 Additional Results and Discussions

### 5.1 Heterogeneity

I analyze the the heterogeneous EIP effect across different income groups to show that the treatment effects of EIP may vary according to the different ex-ante income level. In addition, the heterogeneity analysis also helps rule out the possibility that the decrease in human mobility is caused by an increase in leisure due to the positive income shock, as mentioned in the end of the previous section. Recall that when the household’s income level is lower than a threshold, her optimal choice on the mitigation effort may be bounded by the status quo social aggregate level of mitigation effort. Under such circumstance, if EIP amount is not large enough to make her optimal choice unbounded, then EIP has no effect on her mobility. Since the income threshold is unknown and difficult to be determined empirically, I do not apply parametric assumptions. Instead, I analyze the heterogeneity in a non-parametric way by allowing the effect of EIP treatment to be different across arbitrarily defined income groups.

In the DID framework with continuous treatment, I define 5 household income groups: $0−20, 000, $20, 000−40, 000, $40, 000−60, 000, $60, 000−100, 000 and $100, 000−150, 000. Let the fraction of households in each income bin be denoted by 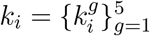, then the continuous treatment is defined as

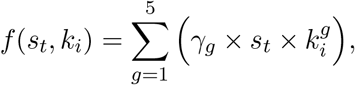

where *γ*_*g*_ is the average treatment effect coefficient of EIP received by income group *g*. Table 3 reports the results. The first 5 variables reflect the amount of EIP received by different income groups. The results indicate that the households with income less than $20,000 do not change their mobility (therefore, do not increase their contribution to the mitigation effort) even after receiving EIP, suggesting that after receiving EIP, these households are still below the income threshold. To confirm that “Income $0 − 20, 000” is a good approximation for the households bounded by the status quo social aggregate mitigation effort after receiving EIP, Table B4 in the appendix shows the immediate next income group, $20, 000−25, 000, has a significantly positive EIP effect. All other income groups increase their public good contribution, which suggests that after receiving EIP, their income exceeds the threshold. The income group $100, 000 − 150, 000 has a relatively smaller estimated effect because of two reasons. First, some households (i.e., households of single) in the group $100, 000 − 150, 000 may receive reduced EIP (less than $1,200).^21^ Second, the mitigation effort is more likely to be the necessities with an concave Engel curve and diminishing slope as the income increases, rather than a luxury good. The coefficients on the other variables in the regression are very similar to the main results.

**Table 3:** Heterogeneous EIP Effect in Income

|  | *Dependent variable:* |  |
| --- | --- | --- |
|  | *Median Home Dwell Time (minutes, in log)* |  |
|  | (State Policy) | (County Policy) |
| EIP, Income \$0-20,000 | −0.0172<br>(0.0116) | 0.0055<br>(0.0133) |
| EIP, Income \$20,000-40,000 | 0.0646***<br>(0.0135) | 0.0824***<br>(0.0142) |
| EIP, Income \$40,000-60,000 | 0.0610***<br>(0.0136) | 0.0688***<br>(0.0156) |
| EIP, Income \$60,000-100,000 | 0.0777***<br>(0.0099) | 0.0752***<br>(0.0117) |
| EIP, Income \$100,000-150,000 | 0.0497***<br>(0.0108) | 0.0350***<br>(0.0131) |
| County COVID-19 Confirmed Case (per 100) | 0.0003*<br>(0.0002) | 0.0002<br>(0.0002) |
| County COVID-19 Death (per 100) | −0.0041***<br>(0.0012) | −0.0035***<br>(0.0012) |
| Social Distancing (State/County) | 0.0093*<br>(0.0053) | 0.0131<br>(0.0096) |
| Non-essential Services Closure (State/County) | 0.0046<br>(0.0088) | −0.0134<br>(0.0146) |
| Shelter-in-Place (State/County) | 0.0039<br>(0.0088) | 0.0024<br>(0.0131) |
| Block Group FE | Y | Y |
| Date FE | Y | Y |
| Observations | 4,317,586 | 2,541,943 |
| R<sup>2</sup> | 0.8682 | 0.8769 |
| Adjusted R<sup>2</sup> | 0.8612 | 0.8704 |
| Residual Std. Error | 0.4010 (df = 4101488) | 0.4006 (df = 2414698) |

Note: \*p<0.1; \*\*p<0.05; \*\*\*p<0.01
Standard errors are clustered at state level.

Next, I use the event study to reaffirm the findings in the DID. I follow the same setting in Section 4.2 to compare the daily “Median Home Dwell Time” between the high and low income CBGs. In further, with the prior that “Income $0 − 20, 000” is a good approximation for the households with bounded optimal mobility after receiving EIP, I divide the low income CBGs into two subgroups: the bounded CBGs and the unbounded CBGs, depending on whether there are majority of the households with income less than $20, 000 (according to DID results above) or not. If “Income $0 − 20, 000” is a good approximation for the bounded households, then we should expect much smaller EIP treatment effect for the bounded CBGs than the unbounded CBGs. Consider the following regression:

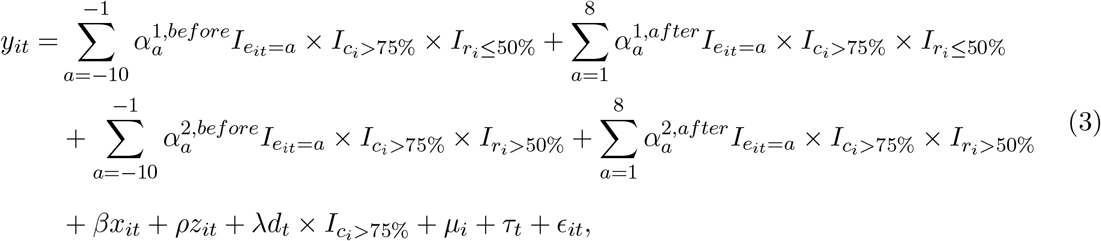

where *r*_*i*_ is the share of households with income less than $20, 000, 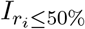 and 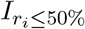 are indicator functions. 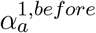 and 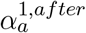 capture the daily gap between the high income CBGs and the unbounded low income CBGs in which less than half of the households have income lower than $20, 000; 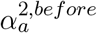 and 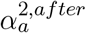 capture the daily gap between the high income CBGs and the bounded low income CBGs in which more than half of the households have income lower than $20, 000. Using the state and county policy data, Figure 5a and 6a plot the results for estimated 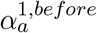 and 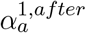, Figure 5b and 6b plot the results for estimated 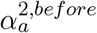 and 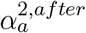. We find that the estimated daily gap between the high income CBGs and the unbounded low income CBGs plotted in Figure 5a and 6a follows the similar patterns as the estimated daily gap between the high and low income CBGs shown in Figure 3 and 4, suggesting that there is a strong EIP treatment effect on the human mobility in the unbounded CBGs. However, according to Figure 5b and 6b, there is no clear evidence that the EIP treatment affects the human mobility in the bounded low income CBGs. These results are consistent with the results from DID analysis of heterogeneous effects and support the argument that “Income $0 − 20, 000” is a good approximation for the households with bounded optimal mobility even after receiving EIP.

**Figure 5:**
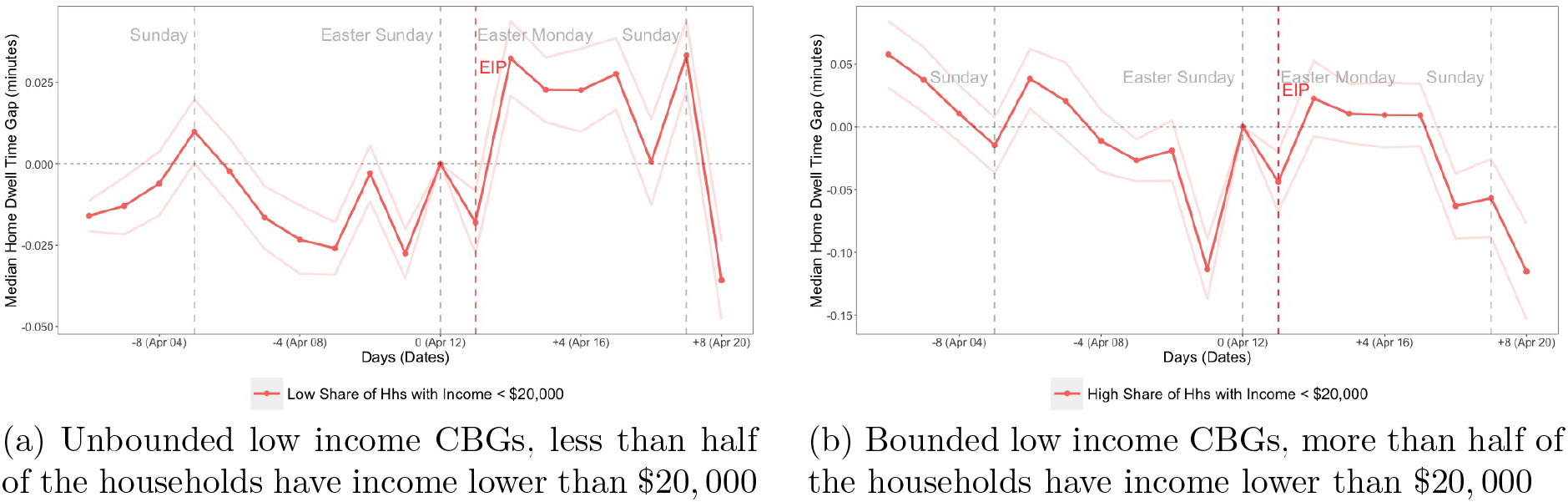
Event Study of Heterogeneous EIP Effect, State Policies

**Figure 6:**
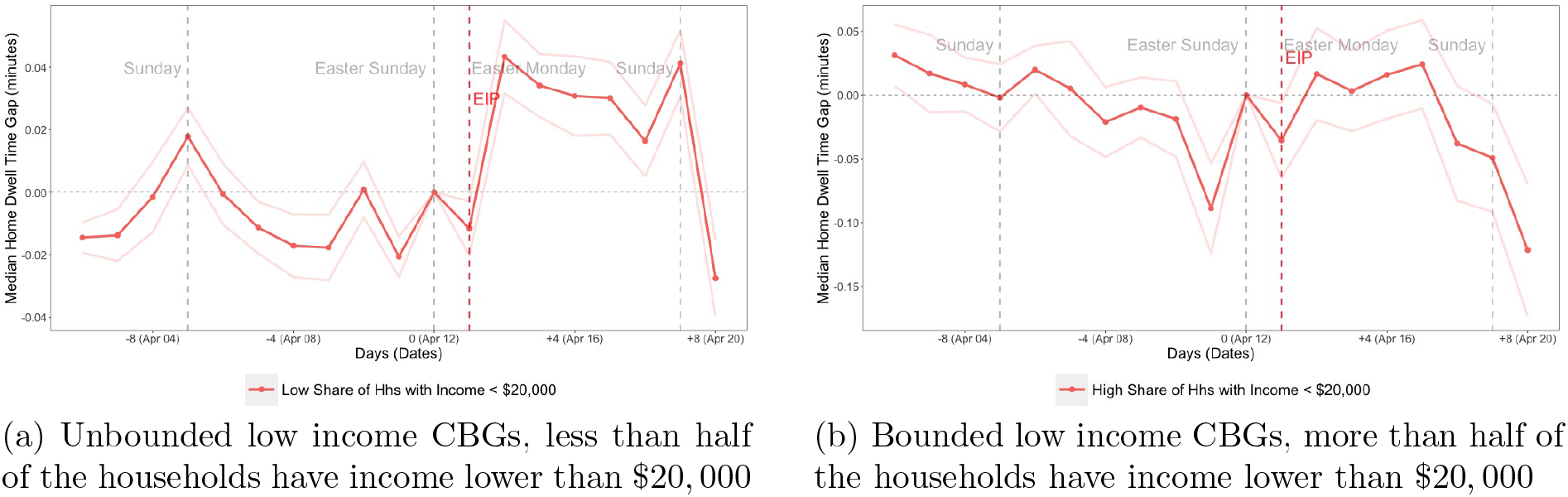
Event Study of Heterogeneous EIP Effect, County Policies

There is another concern about the results of the heterogeneous analysis, which is related to the timing of receiving EIP. In fact, all the direct deposits were received in the week of April 13, 2020, but for the households who do not have the direct deposits, the paper checks arrived by mail in the subsequent weeks. My analysis only captures the EIP effect through the direct deposits, so it is a lower bound of the true EIP effect on human mobility. If the low income households are less likely to have the direct deposit accounts, that could also explain the null effect of EIP shock for these households. On the other hand, the sign and significance of the estimate coefficients are reliable as long as the number of households without direct deposits does not dominate the whole population.

### 5.2 Robustness Check

In order to enhance the reliability of the empirical results, I conduct several robustness checks based on the DID framework with continuous treatment. The first robustness exercise is a placebo test. I assign a placebo treatment in the pre-treatment period between April 1st, 2020 and April 12th, 2020. Since the actual EIP treatment period is about half the length of the pre-treatment period, I assign the similar placebo treatment from April 9th, 2020 to April 12th, 2020. The first panel of Table 4 reports the results. I do not find a statistically significant coefficient on the placebo treatment in the sample with state-level policy controls, though in the sample with the county-level policy the placebo treatment is positive at 10% level. These results favor the causal inference that EIP significantly increases the home dwell time.^22^

**Table 4:** Robustness Checks

| *Panel I: Placebo Test* |  |  |
| --- | --- | --- |
|  | *Dependent variable:* |  |
|  | *Median Home Dwell Time (minutes, in log)*<br>(State Policy) | (County Policy) |
| Placebo Treatment (April 9th - April 12th) | 0.0069<br>(0.0090) | 0.0167*<br>(0.0094) |
| County COVID-19 Confirmed Case (per 100) | 0.0005*<br>(0.0003) | 0.0006**<br>(0.0003) |
| County COVID-19 Death (per 100) | -0.0069***<br>(0.0023) | -0.0074***<br>(0.0020) |
| Social Distancing (State/County) | 0.0235***<br>(0.0062) | 0.0082<br>(0.0200) |
| Non-essential Services Closure (State/County) | 0.0098<br>(0.0064) | -0.0097<br>(0.0141) |
| Shelter-in-Place (State/County) | 0.0114<br>(0.0082) | 0.0055<br>(0.0125) |
| Block Group FE | Y | Y |
| Date FE | Y | Y |
| Observations | 2,591,077 | 1,525,503 |
| R<sup>2</sup> | 0.8776 | 0.8856 |
| Adjusted R<sup>2</sup> | 0.8664 | 0.8752 |
| Residual Std. Error | 0.3815 (df = 2375000) | 0.3827 (df = 1398276) |
| *Panel II: Allowing State/County Specific Time Trend* |  |  |
|  | *Dependent variable:* |  |
|  | *Median Home Dwell Time (minutes, in log)*<br>(State Specific Time Trend) | (County Specific Time Trend) |
| Economic Impact Payment | 0.0293***<br>(0.0075) | 0.0521***<br>(0.0087) |
| County COVID-19 Confirmed Case (per 100) | 0.0002***<br>(0.0000) |  |
| County COVID-19 Death (per 100) | -0.0033***<br>(0.0003) |  |
| Block Group FE | Y | Y |
| Date FE × County/State FE | Y | Y |
| Observations | 4,317,586 | 2,541,943 |
| R<sup>2</sup> | 0.8687 | 0.8780 |
| Adjusted R<sup>2</sup> | 0.8618 | 0.8712 |
| Residual Std. Error | 0.4002 (df = 4100545) | 0.3994 (df = 2408437) |

*Note:* \*p<0.1; \*\*p<0.05; \*\*\*p<0.01 Standard errors are clustered at state level.

Second, I test the sensitivity of the estimated average EIP treatment effect on a relaxed time trend assumption. In the DID framework, the causal inference relies on the common time trend assumption. However, with continuous treatment, there is no clear distinction between the treatment and control groups, so it is not feasible to estimate daily gap between two groups in the pre-treatment period to verify the common time trend. Alternatively, the causal relationship can be inferred by a more relaxed assumption: the unobserved time-varying variables nested in the residual term are not correlated with both the treatment and the outcome variables. If there exists an unobserved time-varying variable that confounds the treatment, then the results should be sensitive to relaxing the time trend restrictions. In the main analysis, I assume all CBGs follow the same time trend. Here I relax this assumption by allowing state and county specific time fixed effects. The second panel of Table 4 reports the results. Since the state/county specific time fixed effects already account for the temporal variation at the state/county level, the state/county specific time-variant variables are omitted in the regressions. The results are identical to the main results, so we can conclude that there is no unobserved confounders at state and county level that affect the reliability of the estimated average EIP treatment effect in the main analysis.

I also run an additional robustness exercise by considering a different empirical model: the regression discontinuity design with a sharp treatment discontinue point at the EIP treatment starting date. The details are available in the Appendix B.5. The results show that there is a positive and significant upward shift at the discontinue point (the EIP treatment starting date), which is consistent to the main analysis.

## 6 Conclusion

During the 2020 COIVD-19 pandemic, mitigating the virus spreading heavily depends on every individual’s mitigation effort contribution in the society. To increase the individual and social aggregate mitigation effort, a commonly used government instrument is the mandatory policies that set minimum mitigation effort requirement for all individuals. However, in many countries including the United States, due to varies of economic, social and politic issues, it is very difficult to strictly enforce the minimum mitigation effort requirement. In this paper, I propose an argument that other than the mandatory policies, the income compensation policy is an alternative government instrument, which provides a possible decentralized solution to increase the individual and social mitigation effort.

To empirically test whether the income compensation increases the individuals’ mitigation effort contribution, I use the first round EIP in the United States as a quasi-natural experiment, and study the effect of receiving EIP on human mobility (home dwell time), where human mobility (home dwell time) is an indicator of the the COVID-19 mitigation effort. The empirical results show that EIP increases the daily home dwell time by 3% – 5% on average, though this increase is driven by the households with ex-ante income greater than $20,000. For households with income below $20,000, the current EIP amount is insufficient to motivate them to increase the mitigation effort.

By exploring the public good nature of the COVID-19 mitigation effort and the relationship between the individual’s income and mitigation effort contribution, this paper highlights an unintended benefit of EIP: in addition to providing economic assistance during the pandemic, EIP also motivate the individuals to increase their COVID-19 mitigation effort. However, there are limitations in this paper. First, the current empirical analysis is at CBG aggregate level instead of individual level, because the lack of the individual level data of EIP reception. In order to better understanding the individual behavior on the COVID-19 mitigation, further analysis at individual level is required. Second, due to the limited access to Safegraph human mobility data, the data period in this paper is restricted within 2020, which makes the control group unavailable in the analysis since almost all the CBGs received EIP. In the further study, in order to provide more robust empirical evidences, it is worth to collect the data for previous years as the control group and conduct a cross-year analysis.

## Data Availability

All the data except human mobility data are publicly available. To obtain human mobility data, please contact Safegraph.inc

## A Mathematic Appendix

### A.1 Theoretical Model

I construct a theoretical model to describes how the change in income affects the individual’s behavior in voluntarily contributing public good, and fit the model to the COVID-19 pandemic to explain why receiving EIP could potentially increase the voluntary contribution to the pandemic mitigation effort. In specific, I assume the public good is a normal good, so generally we could expect an increase in income to raise voluntary public good contribution. However, given the normal good properties, the individuals with relatively lower income would prefer smaller amount of public good than the higher income individuals. Since the public good is non-rival and non-excludable, with sufficiently low income, individuals face over-supply of the public good that is available in the society that is voluntarily contributed by the other individuals with higher income. Therefore, these lower income individuals have zero motivation to voluntarily contribute, and a marginal increase in income does not change their voluntary contribution to public good, which remains zero.

The model follows the setting of Kotchen and Moore (2007). Assume there are *N* individuals in the economy indexed by *i* = 1, 2, …, *N*. Let *y*_*i*_ be the individual income, *y*_*i*_ is pre-determined and assumed to be constant across the study period^23^. The aggregate public good contribution *G* can be divided into two parts: the mandatory contribution level determined by government policies 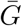, and the voluntary contribution *G*′. Individuals solve the following utility maximization problem:

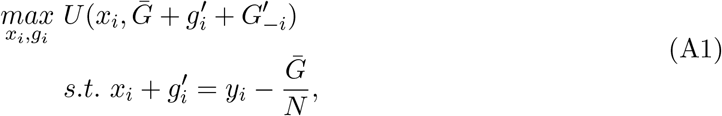

where *x*_*i*_ is the numeraire consumption good, 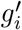 is the voluntary contribution from individual *i*, and 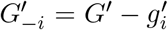 is the aggregate voluntary contribution from others. 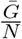 is the per capita minimum contribution to the public good and is subtracted from the individual’s income, *y*_*i*_, to determine the disposable income. I assume that both the numeraire good and the public good are normal goods, so that there exists an unique Nash equilibrium public good contribution (Bergstrom et al., 1986; Kotchen and Moore, 2007). Solving the utility maximization problems with public goods is challenging because no individual is able to fully determine the public good level. I simplify the problem by translating it into an equivalent problem with two private goods and an additional constraint that preserves the public good properties:

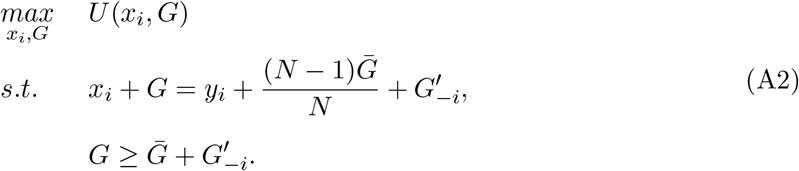

To illustrate the equivalence between two maximization problems, 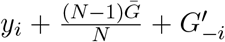 can be interpreted as the individual *i*’s hypothetical income, as if she is endowed with the economy-wide contribution to the public good, so hypothetically the individual is able to decide the total public good contribution level *G*. However, individual *i* is only able to decide her own voluntary public good contribution 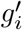, so her choice of *G* is strictly bounded by the constraint 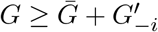

Solving the utility maximization problem yields the following solution:

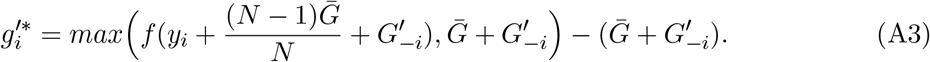

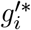 is individual *i*’s optimal voluntary contribution to the public good. The first term of equation A3 is illustrated by the red dotted line in Figure A1, which can be interpreted as the total public good contribution chosen by individual *i*, who is endowed with the hypothetical income, and her choice is bounded by the mandatory requirement plus the voluntary contribution from others 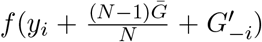 is illustrated by the blue dash line in Figure A1, which is the Engel curve of the public good contribution as a function of the hypothetical income, ignoring the constraint of the minimum required contribution. With the assumption that both the numeraire good and the public good are normal goods, the first derivative of the Engel curve is 0 < *f*′(·) < 1.^24^ As shown in Figure A1, if individual *i* is endowed with the hypothetical income, and her optimal total public good contribution 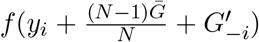 is not bounded by the constraint, then 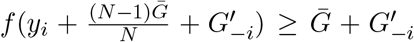 and individual *i* has non-negative voluntary public good contribution. If 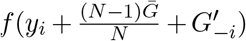 is bounded by the constraint, then 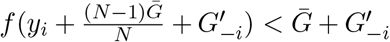 and individual *i* has zero voluntary public good contribution.

Let 𝔾^∗^ be the unique aggregate contribution to the public good in equilibrium. If 𝔾^∗^ is bounded by the mandatory requirement 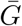, then 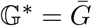, and each individual only contributes at the mandatory level 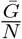. This implies 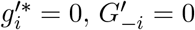, and 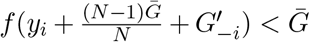 for all *i* ∈ {1, …, *N*}. It can be shown that

#### Proposition 1

*If* 𝔾^∗^ *is bounded by the mandatory requirement* 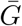, *then there exists an upper bound income level* 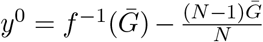 *so that* 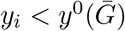 *for all i. A higher* 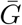 *yields higher* 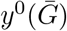.^25^

If 𝔾^∗^ is not bounded by the mandatory requirement 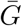, then 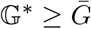. There exists a subset of individuals among the population, denoted by *I*^∗^, so that the optimal choice for every individual in the subset *I*^∗^ is not bounded by the constraint. For each individual *i* ∈ *I*^∗^, we have 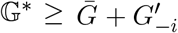 and 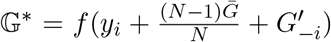. Individual *i*’s voluntary contribution to the public good becomes

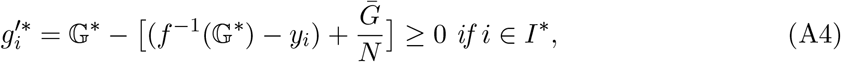

where 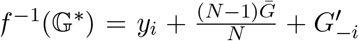, and 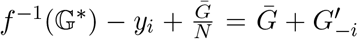 is the summation of the public good contribution from others (both mandatory and voluntary) and the mandatory public good contribution from individual *i*. We can prove that

#### Proposition 2

*If* 𝔾^∗^ *is not bounded by the mandatory requirement* 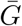, *then there exist income threshold* 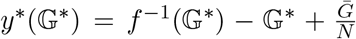, *so that y*_*i*_ ≥ *y*^∗^(𝔾^∗^) *for each individual i* ∈ *I*^∗^ *and y*_*i*_ *< y*^∗^(𝔾^∗^) *for each individual i* ∉ *I*\*. *The higher* 𝔾^∗^ *yields higher y*^∗^(𝔾^∗^).^26^

It is easy to show that 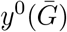 is the lower bound of *y*^∗^(𝔾^∗^) when 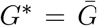.^27^ The aggregate contribution to the public good is 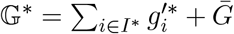, which satisfies

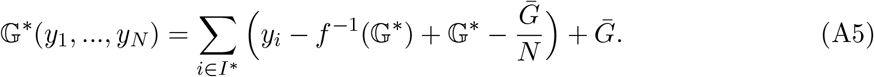

Taking the partial derivative of equation A5 with respect to *y*_*i*_ yields the following result:

#### Proposition 3

*If* 𝔾^∗^ *is not bounded by the mandatory requirement* 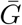, *then we have* 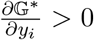 *if i* ∈ *I*^∗^,and 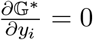.^28^

**Figure A1:**
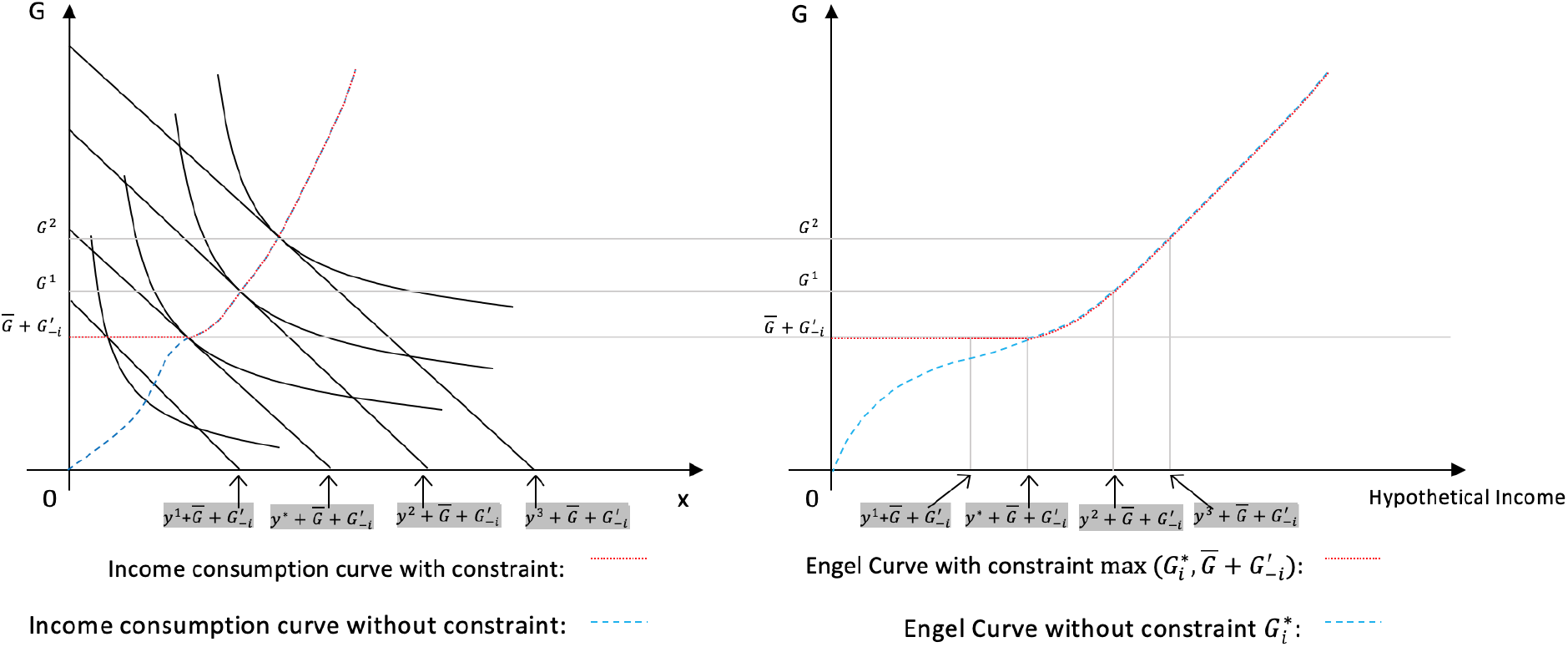
Engel curve with/without the constraint

With a strictly positive mandatory policy 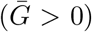, the propositions derived above are not exclusive to the public good. Individual’s purchase of private normal goods has similar properties in response to an positive income shock. However, if there is no mandatory requirement of public good contribution, or the government cannot enforce their policy effectively, then 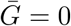. In this case, it is easy to show that the proposition 2 and 3 still hold exclusively only for the public good: although an individual’s optimal choice of aggregate public good is no longer bounded by the government mandatory level, it may still be bounded by the others’ choices such that 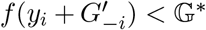. By setting 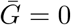, all the results and proofs related to proposition 2 and 3 are still valid.

In the context of the COVID-19 pandemic, Propositions 1-3 lead to the following policy implications. The government has two instruments to raise the social contribution to the public good: enforce a mandatory contribution requirement 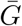; or relax the individuals’ budget constraint. As described previously, if the government set an enforceable mandatory contribution requirement 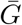, the government may face two possible scenarios: (i) the aggregate contribution to the public good is bounded by the mandatory contribution requirement; (ii) the aggregate contribution to the public good is not bounded by the mandatory contribution requirement. In the first scenario, proposition 1 and 2 imply that a marginal increase in the individuals’ income does not yield an increase in the public good contribution. One must raise at least one arbitrary individual’s income until it exceeds the upper bound 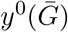, so that a further marginal increase in the same individual’s income will motivate her to voluntarily contribute additional amounts of the public goods beyond the mandatory level, and the aggregate public good contribution increases. In the second scenario, proposition 2 and 3 imply that a marginal increase in the individuals’ income yields an increase in the public good contribution only if the individuals’ original income is higher than the threshold *y*^∗^(𝔾^∗^). If it is hard for the government to enforce a mandatory contribution requirement, or the enforcement is ineffective, then 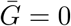 and same implications can be concluded as described in the second scenario above: the marginal income compensation should be properly targeted to the individuals with original income higher than the threshold, or the amount of income compensation should be carefully customized to make the individuals with low income to exceed the threshold.

### A.2 Proof of Proposition 1

According to equation A3, if the equilibrium voluntary consumption of the public good is zero, then 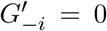 and 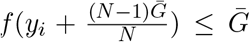 for all *i* ∈ {1, …, *N*}. After inverting *f* (·), we can calculate the upper bound of income under the situation of zero voluntary consumption of the public good:

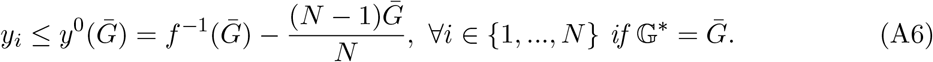

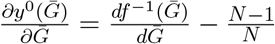. Because 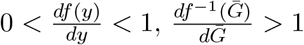, so 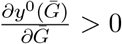.

### A.3 Proof of Proposition 2

For each individual *i* ∈ *I*^∗^, we can calculate the optimal voluntary consumption of the public good by inverting 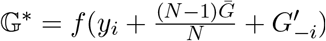:

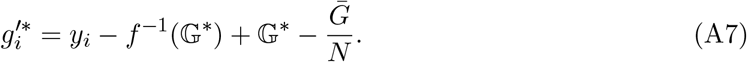

Therefore, the income threshold can be defined as

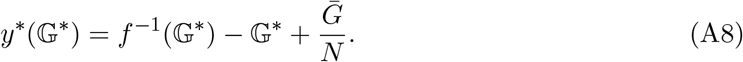

For each individual *i* ∈ *I*^∗^, *y*_*i*_ ≥ *y*^∗^(𝔾^∗^). For each individual *i* ∉ *I*^∗^, 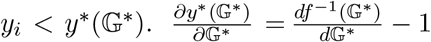. Because Because 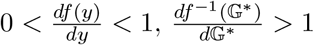, so 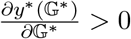.

### A.4 Proof of Proposition 3

Taking the partial derivative of equation A5 yields

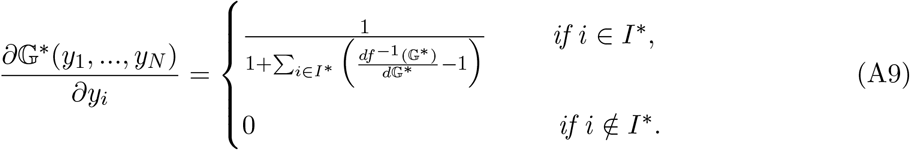

Because 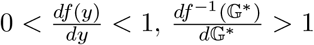. Therefore, 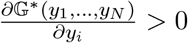 if *i* ∈ *I*\*.

### A.5 Identifying Sample Average Treatment Effect

To properly interpret the treatment coefficient *γ* that is reported as the treatment effect in the regression results tables, I use the following potential outcome framework. Let *i* = 1, 2, …, *N* be the index of all CBGs in the sample. Assume that for any CBG *i* at time *t, y*_*it*_(0) = *µ*_*i*_ + *τ*_*t*_ is the potential outcome if not treated and *y*_*it*_(1) = *γ* + *µ*_*i*_ + *τ*_*t*_ is the potential outcome if CBG *i* is treated. *γ* is the sample average treatment effect (ATE). In the context of this paper, *y*_*it*_(0) is the potential outcome of human mobility if none of the households in CBG *i* receives EIP, and *y*_*it*_(1) is the potential outcome of human mobility if all of the households in CBG *i* receives EIP. *γ* can be interpreted as the sample average increase in “Median Home Dwell Time” of CBGs if all households received an universal EIP. In the data, both *y*_*it*_(0) and *y*_*it*_(1) are hard to observe because for most of CBGs the share of EIP eligible households are neither 0% nor 100%. Therefore, according to the functional form assumption of continuous treatment, most of CBGs receive partial treatment *γk*_*i*_, depending on the share of EIP eligible households *k*_*i*_. Let *t*_0_ be the pre-treatment days and *t*_1_ be the treatment days, in the data, if we observe 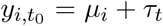 and 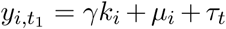 The ATE can be identified by comparing any pair of CBGs {*i, j*} with *i* ≠ *j*:

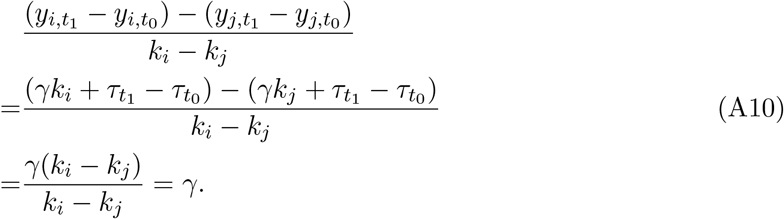

OLS estimates 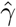 gives a weighted average of all these comparisons. A rigorous proof is omitted since it is beyond the scope of this paper.

Fricke (2017) raised an concern about using Difference-in-Differences to estimate the average treatment effect when there are multiple treatments. He found that Difference-in-Differences identifies a lower bound of average treatment effect if multiple treatments are distributed endogenously in the treatment group. In this paper, CBGs receive a single treatment but at different extent, depending on the CBG household income. If the CBG household income is not endogenous to human mobility in the short run at daily basis, then we can conclude that the size of the EIP treatment is exogenous and we do not have the estimation problem described in Fricke (2017).

## B Additional Regression Results

### B.1 Placebo test in February and March

I run additional robustness analysis to reaffirm the findings of this paper. In Table B1, the placebo tests are done by using different samples with the same days from February and March. The results show that the coefficients of the placebo treatment are insignificant for February sample. However, the coefficients of the placebo treatment is significantly negative for March sample, probably because the major wave of lay-off/furlough in late March disproportionately creates larger negative income shock on the lower income families, so that they reduce more mitigation effort contribution in response.

**Table B1:**
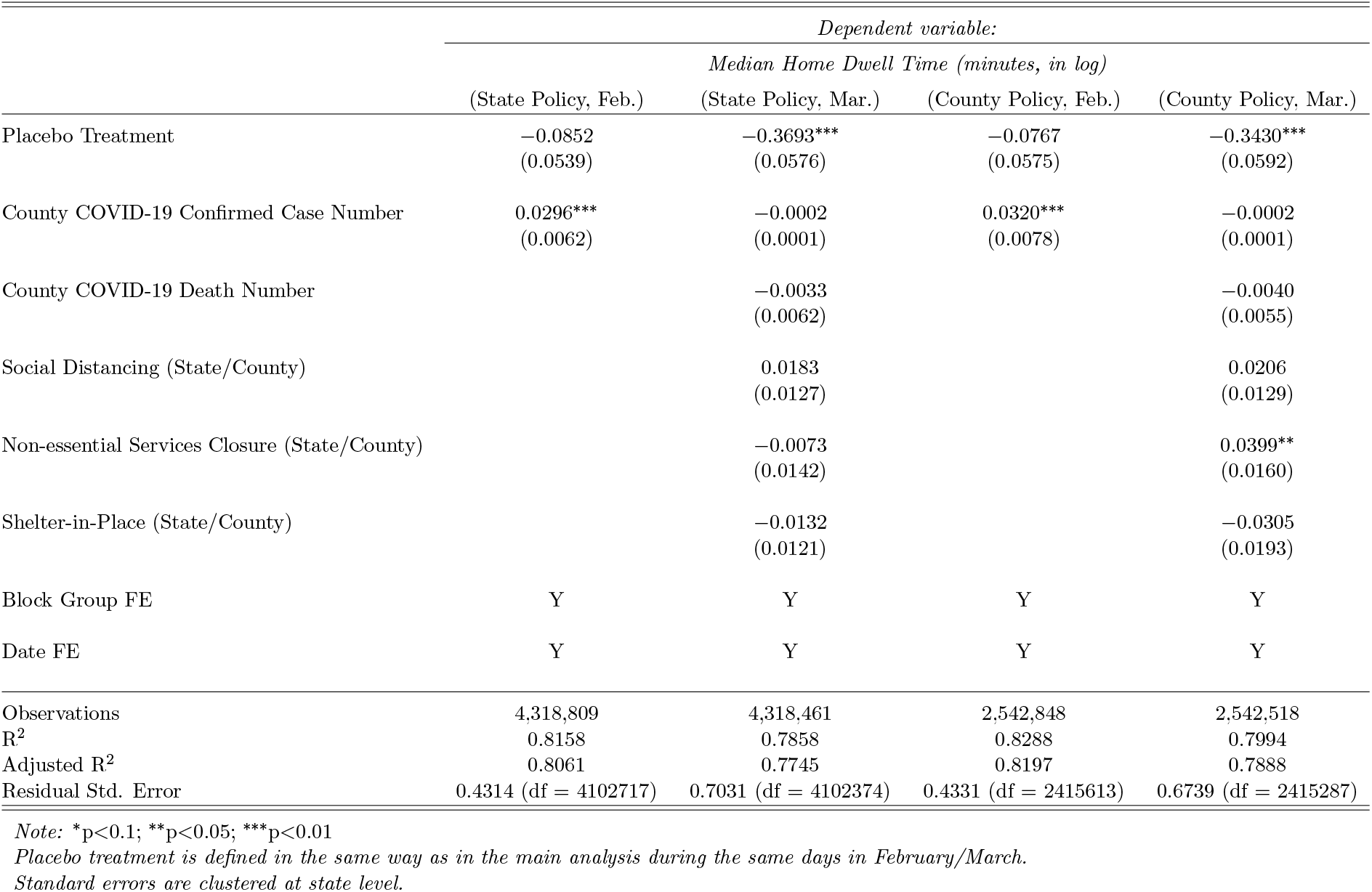
Robustness Check: Placebo Test on Previous Months

|  | *Dependent variable:* |  |  |  |
| --- | --- | --- | --- | --- |
|  | *Median Home Dwell Time (minutes, in log)* |  |  |  |
|  | (State Policy, Feb.) | (State Policy, Mar.) | (County Policy, Feb.) | (County Policy, Mar.) |
| Placebo Treatment | -0.0852<br>(0.0539) | -0.3693***<br>(0.0576) | -0.0767<br>(0.0575) | -0.3430***<br>(0.0592) |
| County COVID-19 Confirmed Case Number | 0.0296***<br>(0.0062) | -0.0002<br>(0.0001) | 0.0320***<br>(0.0078) | -0.0002<br>(0.0001) |
| County COVID-19 Death Number |  | -0.0033<br>(0.0062) |  | -0.0040<br>(0.0055) |
| Social Distancing (State/County) |  | 0.0183<br>(0.0127) |  | 0.0206<br>(0.0129) |
| Non-essential Services Closure (State/County) |  | -0.0073<br>(0.0142) |  | 0.0399**<br>(0.0160) |
| Shelter-in-Place (State/County) |  | -0.0132<br>(0.0121) |  | -0.0305<br>(0.0193) |
| Block Group FE | Y | Y | Y | Y |
| Date FE | Y | Y | Y | Y |
| Observations | 4,318,809 | 4,318,461 | 2,542,848 | 2,542,518 |
| R<sup>2</sup> | 0.8158 | 0.7858 | 0.8288 | 0.7994 |
| Adjusted R<sup>2</sup> | 0.8061 | 0.7745 | 0.8197 | 0.7888 |
| Residual Std. Error | 0.4314 (df = 4102717) | 0.7031 (df = 4102374) | 0.4331 (df = 2415613) | 0.6739 (df = 2415287) |

Note: \*p<0.1; \*\*p<0.05; \*\*\*p<0.01
Placebo treatment is defined in the same way as in the main analysis during the same days in February/March.
Standard errors are clustered at state level.

### B.2 Policy Effect

In this analysis, I include the interaction between the NPI policies and the share of EIP eligibility households in regression equation 2 to test if the main results is contaminated by the difference in policy effects between EIP eligible and non-eligible households. As expected, most coefficients of interaction terms and policy dummies are insignificant, and EIP coefficients remain significantly positive with similar magnitude as in the main results.^29^

**Table B2:**
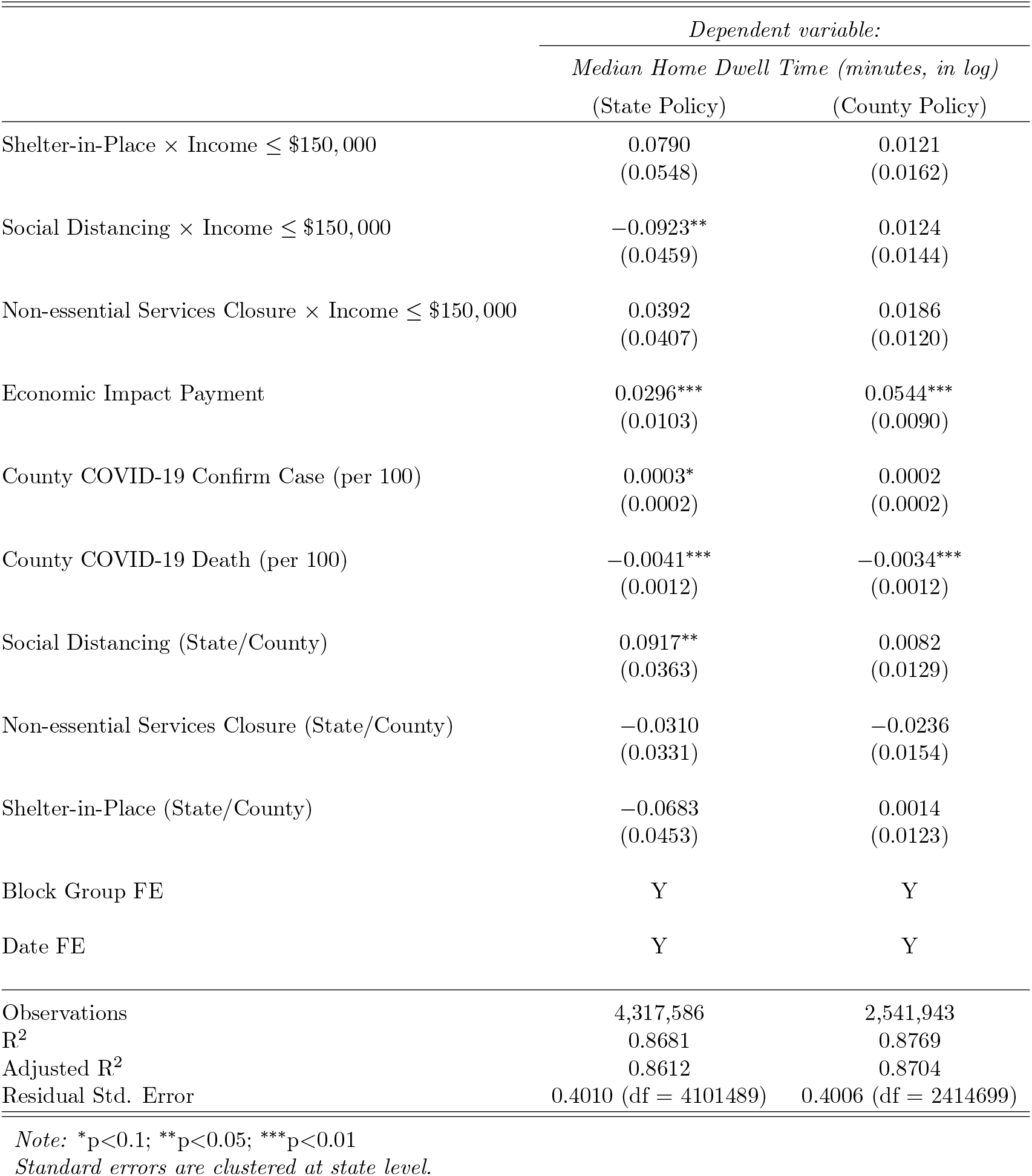
Policy Effects on Human Mobility

|  | *Dependent variable:* |  |
| --- | --- | --- |
|  | *Median Home Dwell Time (minutes, in log)* |  |
|  | (State Policy) | (County Policy) |
| Shelter-in-Place × Income ≤ \$150,000 | 0.0790<br>(0.0548) | 0.0121<br>(0.0162) |
| Social Distancing × Income ≤ \$150,000 | -0.0923**<br>(0.0459) | 0.0124<br>(0.0144) |
| Non-essential Services Closure × Income ≤ \$150,000 | 0.0392<br>(0.0407) | 0.0186<br>(0.0120) |
| Economic Impact Payment | 0.0296***<br>(0.0103) | 0.0544***<br>(0.0090) |
| County COVID-19 Confirm Case (per 100) | 0.0003*<br>(0.0002) | 0.0002<br>(0.0002) |
| County COVID-19 Death (per 100) | -0.0041***<br>(0.0012) | -0.0034***<br>(0.0012) |
| Social Distancing (State/County) | 0.0917**<br>(0.0363) | 0.0082<br>(0.0129) |
| Non-essential Services Closure (State/County) | -0.0310<br>(0.0331) | -0.0236<br>(0.0154) |
| Shelter-in-Place (State/County) | -0.0683<br>(0.0453) | 0.0014<br>(0.0123) |
| Block Group FE | Y | Y |
| Date FE | Y | Y |
| Observations | 4,317,586 | 2,541,943 |
| R<sup>2</sup> | 0.8681 | 0.8769 |
| Adjusted R<sup>2</sup> | 0.8612 | 0.8704 |
| Residual Std. Error | 0.4010 (df = 4101489) | 0.4006 (df = 2414699) |

*Note:* \*p<0.1; \*\*p<0.05; \*\*\*p<0.01 Standard errors are clustered at state level.
<sup>29</sup>The coefficients of social distancing policy is significant in state policy sample. Further study is desired to explain.

### B.3 Heterogeneous EIP Effect with Relaxed Time Trend Assumption

Table B3 reports the results of heterogeneous EIP effect under the relaxed time trend assumptions. I find the coefficients are identical with Table 3, which in further increases the credibility of the assumption that EIP treatments are not confounded by other unobserved time-variant variables.

**Table B3:**
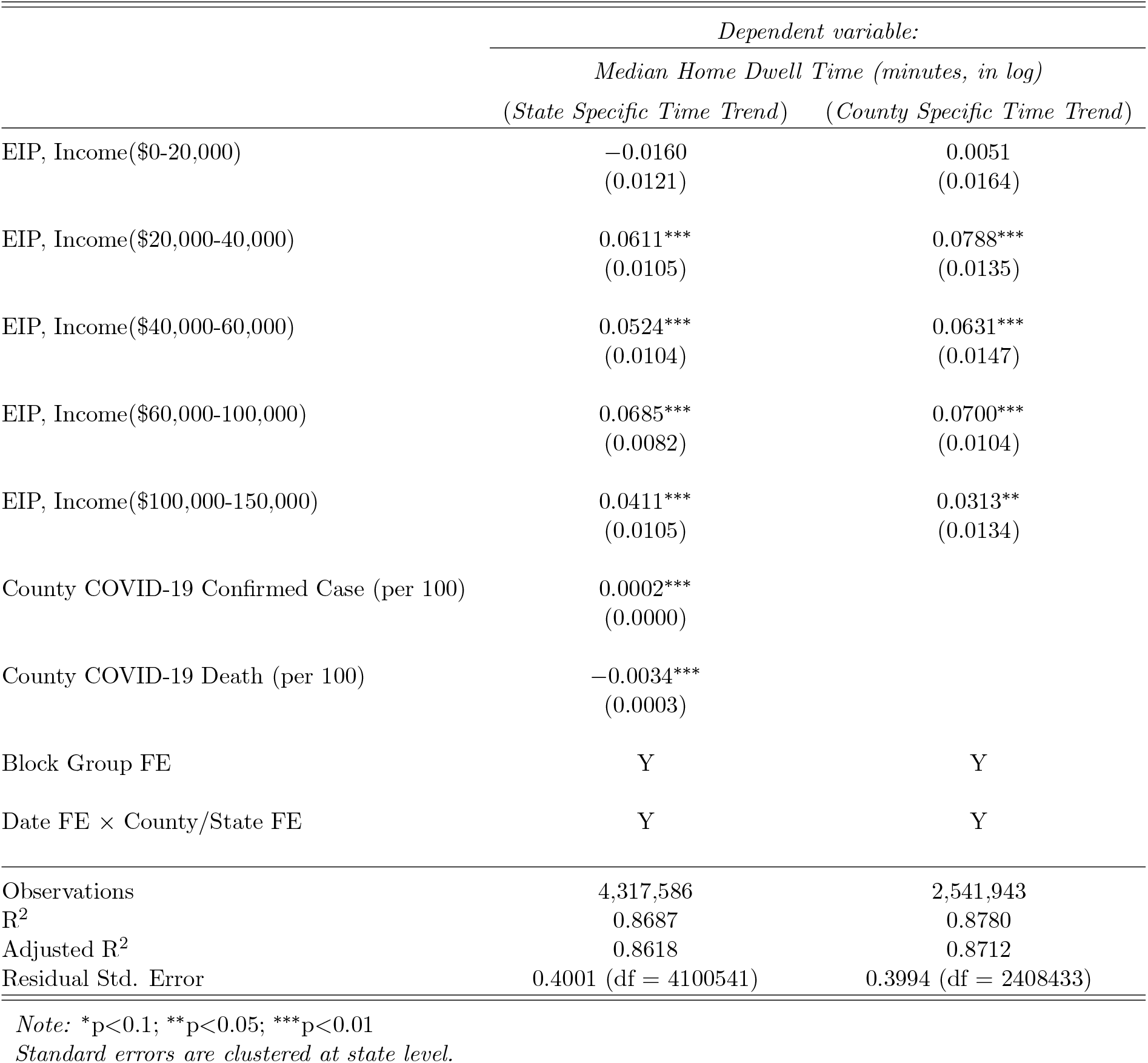
Robustness Check: Heterogeneous EIP Effect in Income, Allowing State/County Specific Time Trend

### B.4 Alternative Division of Income Groups

**Table B4:** Heterogeneous EIP Effect in Income, Identify the Income Threshold

|  | *Dependent variable:* |  |
| --- | --- | --- |
|  | *Median Home Dwell Time (minutes, in log)* |  |
|  | (State Policy) | (County Policy) |
| EIP, Income \$0-20,000 | -0.0171<br>(0.0115) | 0.0047<br>(0.0132) |
| **EIP, Income \$20,000-25,000** | **0.0632\*\*\***<br>(0.0214) | **0.1086\*\*\***<br>(0.0251) |
| EIP, Income \$25,000-40,000 | 0.0651***<br>(0.0136) | 0.0731***<br>(0.0143) |
| EIP, Income \$40,000-60,000 | 0.0610***<br>(0.0136) | 0.0693***<br>(0.0156) |
| EIP, Income \$60,000-100,000 | 0.0776***<br>(0.0099) | 0.0756***<br>(0.0117) |
| EIP, Income \$100,000-150,000 | 0.0497***<br>(0.0107) | 0.0353***<br>(0.0132) |
| County COVID-19 Confirm Case (per 100) | 0.0003*<br>(0.0002) | 0.0002<br>(0.0002) |
| County COVID-19 Death (per 100) | -0.0041***<br>(0.0012) | -0.0035***<br>(0.0012) |
| Social Distancing (State/County) | 0.0093*<br>(0.0053) | 0.0131<br>(0.0096) |
| Non-essential Services Closure (State/County) | 0.0046<br>(0.0088) | -0.0134<br>(0.0146) |
| Shelter-in-Place (State/County) | 0.0039<br>(0.0088) | 0.0024<br>(0.0131) |
| Block Group FE | Y | Y |
| Date FE | Y | Y |
| Observations | 4,317,586 | 2,541,943 |
| R<sup>2</sup> | 0.8682 | 0.8769 |
| Adjusted R<sup>2</sup> | 0.8612 | 0.8704 |
| Residual Std. Error | 0.4010 (df = 4101487) | 0.4006 (df = 2414697) |

*Note:* \*p<0.1; \*\*p<0.05; \*\*\*p<0.01 Standard errors are clustered at state level.

**Table B5:** Heterogeneous EIP Effect in Income, Alternative Income Groups

|  | *Dependent variable:* |  |
| --- | --- | --- |
|  | *Median Home Dwell Time (minutes, in log)* |  |
|  | (State Policy) | (County Policy) |
| EIP, Income \$0-25,000 | −0.0223*<br>(0.0113) | 0.0070<br>(0.0120) |
| EIP, Income \$25,000-50,000 | 0.0534***<br>(0.0131) | 0.0648***<br>(0.0153) |
| EIP, Income \$50,000-75,000 | 0.0583***<br>(0.0177) | 0.0770***<br>(0.0189) |
| EIP, Income \$75,000-100,000 | 0.0513***<br>(0.0113) | 0.0476***<br>(0.0137) |
| EIP, Income \$100,000-125,000 | 0.0393***<br>(0.0131) | 0.0378***<br>(0.0139) |
| EIP, Income \$125,000-150,000 | 0.0060<br>(0.0177) | −0.0120<br>(0.0241) |
| County COVID-19 Confirm Case (per 100) | 0.0003*<br>(0.0002) | 0.0002<br>(0.0002) |
| County COVID-19 Death (per 100) | −0.0040***<br>(0.0012) | −0.0034***<br>(0.0012) |
| Social Distancing (State/County) | 0.0093*<br>(0.0052) | 0.0134<br>(0.0096) |
| Non-essential Services Closure (State/County) | 0.0047<br>(0.0088) | −0.0134<br>(0.0145) |
| Shelter-in-Place (State/County) | 0.0040<br>(0.0088) | 0.0026<br>(0.0131) |
| Block Group FE | Y | Y |
| Date FE | Y | Y |
| Observations | 4,317,586 | 2,541,943 |
| R<sup>2</sup> | 0.8681 | 0.8769 |
| Adjusted R<sup>2</sup> | 0.8612 | 0.8704 |
| Residual Std. Error | 0.4010 (df = 4101487) | 0.4006 (df = 2414697) |

Note: \*p<0.1; \*\*p<0.05; \*\*\*p<0.01

### B.5 Regression Discontinuity

An alternative model specification is the regression discontinuity design (RDD). Consider the following RDD regression:

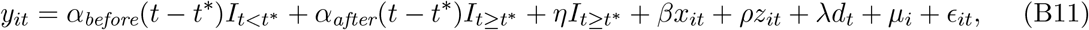

where *t*^∗^ is April 13th, 2020 is the discontinuity cutoff. *I*_*t*<*t*\*_ and *I*_*t*≥*t*\*_ are indicator functions, which equals 1 if its subscript condition is satisfied and 0 otherwise. *α*_*before*_ and *α*_*after*_ are the coefficients of the potentially different slopes at either side of the cutoff. *d*_*t*_ is a dummy variable with *d*_*t*_ = 0 if *t* is a weekday and *d*_*t*_ = 1 if *t* is Saturday or Sunday. *η* is the coefficient of interest, measuring the size of discontinuity at the cutoff date.

Table B6 and B7 report the results using state and county level policy data. In both tables, the first column uses the full sample. It shows that there is a positive and significant upward shift at the discontinuity, suggesting EIP increase home dwell time. This is consistent with the main results where I also find that EIP has a positive effect on human mobility.

To explore the potential heterogeneous EIP effects on households with different income, in the other columns of two tables, I use sub-samples of CBGs selected by the share of low income households to run the regression discontinuity. I define “Low Income” households as these with annual income less than $60,000.^30^ For both tables, the second column uses a sub-sample of CBGs in which more than 75% of households have annual income less than $60,000; the third column uses a sub-sample of CBGs in which the ratio of households with annual income less than $60,000 is between 50% and 75%; the fourth column uses a sub-sample of CBGs in which the ratio of households with annual income less than $60,000 is between 25% and 50%; and the fifth column uses a sub-sample of CBGs in which less than 25% of households have annual income less than $60,000.

According to the heterogeneity results in column 2 – 5 of both tables, I find that for all sub-samples, there is an upward shift at the discontinuity cutoff. Moreover, the CBGs with higher share of low income households have larger upward shift at discontinuity cutoff (0.1321 for state policy data and 0.1360 for county policy data). As the share of low income households decreases, the magnitude of the shift at the cutoff also declines. This is consistent with the previous results for heterogeneous EIP effect in income, suggesting that EIP effect is larger for the CBGs with lower income because more individuals are eligible for and have received EIP.

**Table B6:** Regression Discontinuity Results, State Policy

|  | *Dependent variable:* |  |  |  |  |
| --- | --- | --- | --- | --- | --- |
|  | Median Home Dwell Time (minutes, in log) |  |  |  |  |
|  | Full Sample | Low Income > 75% | Low Income 50% ~ 75% | Low Income 25% ~ 50% | Low Income ≤ 25% |
| Discontinuity Cutoff | 0.0883***<br>(0.0083) | 0.1321***<br>(0.0099) | 0.0965***<br>(0.0081) | 0.0742***<br>(0.0071) | 0.0482***<br>(0.0075) |
| Days Before Cutoff | -0.0024***<br>(0.0008) | -0.0096***<br>(0.0010) | -0.0023***<br>(0.0007) | -0.0002<br>(0.0008) | -0.0003<br>(0.0006) |
| Days After Cutoff | -0.0128***<br>(0.0007) | -0.0169***<br>(0.0010) | -0.0135***<br>(0.0008) | -0.0114***<br>(0.0007) | -0.0092***<br>(0.0007) |
| County COVID-19 Confirm Case (per 100) | 0.0002<br>(0.0002) | 0.0011***<br>(0.0002) | 0.0002<br>(0.0002) | 0.00003<br>(0.0001) | 0.0001*<br>(0.0001) |
| County COVID-19 Death (per 100) | -0.0036***<br>(0.0013) | -0.0081***<br>(0.0016) | -0.0027<br>(0.0017) | -0.0027**<br>(0.0011) | -0.0039***<br>(0.0006) |
| Social Distancing (State) | 0.0014<br>(0.0055) | -0.0092<br>(0.0141) | -0.0085<br>(0.0056) | 0.0111<br>(0.0094) | 0.0244***<br>(0.0081) |
| Non-essential Services Closure (State) | -0.0040<br>(0.0067) | 0.0090<br>(0.0072) | -0.0051<br>(0.0070) | -0.0085<br>(0.0079) | 0.0042<br>(0.0117) |
| Shelter-in-Place (State) | -0.0087<br>(0.0062) | 0.0190**<br>(0.0081) | -0.0030<br>(0.0069) | -0.0249***<br>(0.0076) | -0.0101<br>(0.0079) |
| Weekend | 0.1131***<br>(0.0060) | 0.1163***<br>(0.0052) | 0.1358***<br>(0.0050) | 0.1100***<br>(0.0064) | 0.0515***<br>(0.0059) |
| Block Group FE | Y | Y | Y | Y | Y |
| Observations | 4,317,586 | 692,771 | 1,622,386 | 1,439,674 | 562,755 |
| R<sup>2</sup> | 0.8672 | 0.8599 | 0.8511 | 0.8711 | 0.8851 |
| Adjusted R<sup>2</sup> | 0.8602 | 0.8525 | 0.8432 | 0.8643 | 0.8790 |
| Residual Std. Error | 0.4024 (df = 4101508) | 0.5491 (df = 658050) | 0.3904 (df = 1541212) | 0.3494 (df = 1367646) | 0.3436 (df = 534573) |

Note: \*p<0.1; \*\*p<0.05; \*\*\*p<0.01 Standard errors are clustered at state level.

**Table B7:**
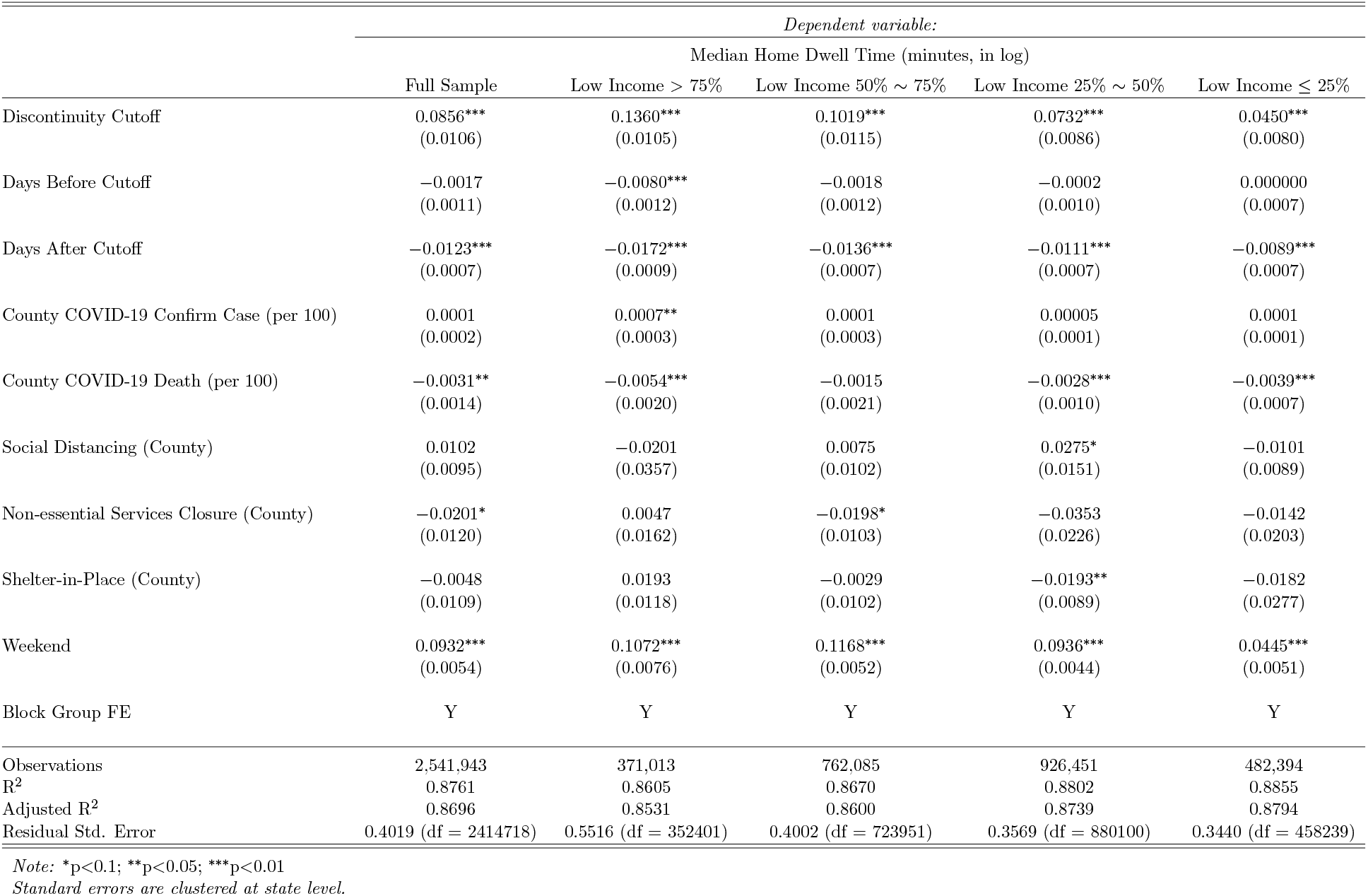
Regression Discontinuity Results, County Policy

## Compliance with Ethical Standards

### Ethical approval

This article does not contain any studies with human participants or animals performed by any of the authors.

## Footnotes

1 It is reasonable to assume that COVID-19 mitigation effort is a normal good, since it is a health related good. Literature usually find the income elasticity of health expenditure positive. For example, Acemoglu et al. (2013) estimated income elasticity of health expenditure equals 0.7 for the United States from 1960-2005. So it is reasonable to assume that the purchase of health related good increases with income.

2 https://home.treasury.gov/policy-issues/cares

3 IRS announced on Twitter that its first EIP deposit occurs on April 11th, 2020. Given it is a Friday, most of EIP direct deposits are more likely to arrive in the week of April 13th. Besides, the expected timeline of EIP published by the House Ways & Means Committee also indicates EIP direct deposit is in the week of April 13th. (https://twitter.com/IRSnews/status/1249062356077944832)

4 See empirical results shown in Table 2 and B2.

5 I use the household income because the census data does not report the individual income distribution.

6 See https://docs.safegraph.com/docs/places-manual.

7 https://github.com/CSSEGISandData/COVID-19/tree/master/csse_covid_19_data/csse_covid_19_time_series

8 The government NPI policy data are collected by Keystone Strategy, partnered with Susan Athey, Stanford Professor of Economics, and Marco Iansiti, Director of Harvard Business School’s Digital Initiative. https://www.keystonestrategy.com/coronavirus-covid19-intervention-dataset-model/

9 Since “Non-essential Services Closure” is always included in the “Shelter-in-Place” order, I assume that the “Non-essential Services Closure” order is substituted by the “Shelter-in-Place” order once the later is issued.

10 2019 income data at CBG level is not available by May, 2020, the time when this research is done.

11 For example, New York State governor avoids using “Shelter-in-Place”, so in the data, New York state has not issued “Shelter-in-Place” order, but only “Non-essential Services Closure”. However, all the counties in New York City have issued “Shelter-in-Place” order.

12 BLS (2020) reports the April unemployment number is 23.1 million, compared to Coibion et al. (2020)’s estimate of over 20 million at the beginning of April. This suggests that the major unemployment wave occurs before the beginning of April, and unemployment rate is relatively stable within the month.

13 Although households with income higher than $150,000 may receive reduced EIP, I exclude them because relative to their ex-ante income, a small lump-sum payment may yield very limited marginal change in their optimal behavior.

14 The threshold of 75% is arbitrarily defined. The pattern remains similar as the threshold varies between 60% and 80%.

15 Dose-response function is frequently used in the medical and environmental literature describing the relationship between exposure to a different level of treatment as a cause and specific outcomes as an effect.

16 There are only 28 CBGs with all households income higher than $150,000.

17 *γ* is estimated and reported in the tables of regression results in the following sectors as the coefficient for “Economic Impact Payment”. The discussion of identifying sample average treatment effect is in the Appendix A.5.

18 See Appendix A.5.

19 COVID-19 death number is the only covariate with significant coefficient for both state and county policy samples.

20 Null effect of the government policies are further discussed in the Appendix B.2.

21 In addition, Table B5 in the appendix shows the regression results for an alternative division of income groups, in which I find the decline in the EIP effect starts from the income group $75, 000 *−* 100, 000, where $75, 000 is the maximum income for single taxpayers to be eligible for EIP.

22 I also run additional placebo tests using different samples with same days from February and March. The results also support the causal inference that EIP significantly increases the home dwell time. See Appendix B.1.

23 The empirical validity of this assumption is discussed later in section 4.1.

24 Let the income level be *y*. Let *f* (*y*) and *h*(*y*) be the Engel curves of the numeraire good *x* and the public good *g. f* (*y*) + *h*(*y*) = *y*. Taking the first derivative yields 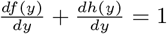. Since both 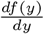 and 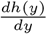 are positive, 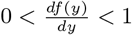.

25 See Appendix A.2 for proofs.

26 See Appendix A.3 for proofs.

27 Because 0 < *f*′(·) < 1, *f*^−1′^(·) > 1, so 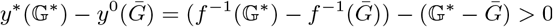.

28 See Appendix A.4 for proofs.

29 The coefficients of social distancing policy is significant in state policy sample. Further study is desired to explain.

30 This is arbitrary defined but close to the national median income level ($64,324) in 2018.

